# Brain ageing in schizophrenia: evidence from 26 international cohorts via the ENIGMA Schizophrenia consortium

**DOI:** 10.1101/2022.01.10.21267840

**Authors:** Constantinos Constantinides, Laura KM Han, Clara Alloza, Linda Antonucci, Celso Arango, Rosa Ayesa-Arriola, Nerisa Banaj, Alessandro Bertolino, Stefan Borgwardt, Jason Bruggemann, Juan Bustillo, Oleg Bykhovski, Vaughan Carr, Stanley Catts, Young-Chul Chung, Benedicto Crespo-Facorro, Covadonga M Díaz-Caneja, Gary Donohoe, Stefan Du Plessis, Jesse Edmond, Stefan Ehrlich, Robin Emsley, Lisa T Eyler, Paola Fuentes-Claramonte, Foivos Georgiadis, Melissa Green, Amalia Guerrero-Pedraza, Minji Ha, Tim Hahn, Frans A Henskens, Laurena Holleran, Stephanie Homan, Philipp Homan, Neda Jahanshad, Joost Janssen, Ellen Ji, Stefan Kaiser, Vasily Kaleda, Minah Kim, Woo-Sung Kim, Matthias Kirschner, Peter Kochunov, Yoo Bin Kwak, Jun Soo Kwon, Irina Lebedeva, Jingyu Liu, Patricia Mitchie, Stijn Michielse, David Mothersill, Bryan Mowry, Víctor Ortiz-García de la Foz, Christos Pantelis, Giulio Pergola, Fabrizio Piras, Edith Pomarol-Clotet, Adrian Preda, Yann Quidé, Paul E Rasser, Kelly Rootes-Murdy, Raymond Salvador, Marina Sangiuliano, Salvador Sarró, Ulrich Schall, André Schmidt, Rodney J Scott, Pierluigi Selvaggi, Kang Sim, Antonin Skoch, Gianfranco Spalletta, Filip Spaniel, Sophia I. Thomopoulos, David Tomecek, Alexander S Tomyshev, Diana Tordesillas-Gutiérrez, Therese van Amelsvoort, Javier Vázquez-Bourgon, Daniela Vecchio, Aristotle Voineskos, Cynthia S Weickert, Thomas Weickert, Paul M Thompson, Lianne Schmaal, Theo GM van Erp, Jessica Turner, James H Cole, Danai Dima, Esther Walton

## Abstract

Schizophrenia (SZ) is associated with an increased risk of life-long cognitive impairments, age-related chronic disease, and premature mortality. We investigated evidence for advanced brain ageing in adult SZ patients, and whether this was associated with clinical characteristics in a prospective meta-analytic study conducted by the ENIGMA Schizophrenia Working Group. The study included data from 26 cohorts worldwide, with a total of 2803 SZ patients (mean age 34.2 years; range 18-72 years; 67% male) and 2598 healthy controls (mean age 33.8 years, range 18-73 years, 55% male). Brain-predicted age was individually estimated using a model trained on independent data based on 68 measures of cortical thickness and surface area, 7 subcortical volumes, lateral ventricular volumes and total intracranial volume, all derived from T1-weighted brain magnetic resonance imaging (MRI) scans. Deviations from a healthy brain ageing trajectory were assessed by the difference between brain-predicted age and chronological age (brain-predicted age difference [brain-PAD]). On average, SZ patients showed a higher brain-PAD of +3.64 years (95% CI: 3.01, 4.26; I^2^ = 55.28%) compared to controls, after adjusting for age and sex (Cohen’s d = 0.50). Among SZ patients, brain-PAD was not associated with specific clinical characteristics (age of onset, duration of illness, symptom severity, or antipsychotic use and dose). This large-scale collaborative study suggests advanced structural brain ageing in SZ. Longitudinal studies of SZ and a range of mental and somatic health outcomes will help to further evaluate the clinical implications of increased brain-PAD and its ability to be influenced by interventions.

## Introduction

Schizophrenia (SZ) is associated with an increased risk of premature mortality, with an average decrease in life expectancy of approximately 15 years [1–3]. This is partially accounted for by suicidal behaviour or accidental deaths, as well as poor somatic health, including cardiovascular and metabolic disease [4–6]. The high prevalence of physical morbidity, long-term cognitive decline, and excess mortality seen in SZ may partly be the result of “accelerated” ageing (i.e., a biological age which “outpaces” chronological age) [7–9]. An increasing number of studies report systemic, age-related biological changes in SZ patients, including elevated levels of oxidative stress, inflammation and cytotoxicity [10, 11]. There is also evidence for progressive brain changes in gray and white matter structures that may begin around or after illness onset [12–18], which may, in part, reflect deviations from normal brain ageing trajectories.

Although chronological age can be predicted accurately with neuroimaging data using machine learning, discrepancies can occur between brain-predicted age (also known as “brain age”) and chronological age [19]. This can be referred to as brain-predicted age difference (brain-PAD). A brain-PAD larger than zero indicates a brain that appears “older” than the person’s chronological age, whereas a brain-PAD lower than zero reflects a “younger” brain than expected at a given chronological age. Higher brain-PAD scores have been associated with a wide range of health-related lifestyle factors and outcomes, including smoking, higher alcohol intake, obesity (or higher BMI), cognitive impairments, major depression, type 2 diabetes, and early mortality [20–25].

To our knowledge, only a few studies have investigated brain age in adults with SZ using various machine learning algorithms or imaging (gray and/or white matter) measures. A higher brain-PAD was consistently shown in SZ patients relative to healthy individuals, with reported scores varying from +2.6 to 7.8 years across studies [26–31]. Furthermore, a greater brain-PAD was observed in first-episode SZ patients [26], and longitudinal data suggests that this gap widens predominantly during the first years after illness onset [29]. As these prior studies were performed with relatively small to moderate sample sizes (range: 43-341 patients), it is important to examine whether brain age findings in SZ can be generalised through large-scale studies consisting of many independent samples worldwide. Two recent mega-analyses with up to 1110 SZ patients across multiple cohorts found a moderate increase in brain-PAD derived from structural T1-weighted MRI (Cohen’s d=0.51) [32] and diffusion tensor imaging (Cohen’s d=0.29) [33], respectively. Validation of those findings, as well as identifying which clinical characteristics or other factors may underlie advanced brain ageing in SZ, could have diagnostic and prognostic implications for patients.

Here, we set out to investigate brain age in over 5000 individuals from the Schizophrenia Working Group within the Enhancing Neuro-Imaging Genetics through Meta-analysis (ENIGMA) consortium (26 cohorts, 15 countries), covering almost the entire adult lifespan (18-73 years). We employed a recently developed multisite brain ageing algorithm based on FreeSurfer-derived gray matter regions of interest (ROIs) [24] to examine brain-PAD differences between SZ patients and healthy controls in a prospective meta-analysis. We hypothesised significantly higher brain-PAD in SZ patients, compared to controls. In addition, we assessed whether a higher brain-PAD in SZ patients was associated with clinical characteristics, such as age of onset, length of illness, symptom severity, and antipsychotic treatment.

## Methods

### Study samples

Twenty-six cohorts from the ENIGMA SZ working group with cross-sectional data from SZ patients (N=2803) and healthy controls (N=2598) were included in this study (18-73 years of age). Details of demographics, location, clinical characteristics (including methods for data harmonization), and inclusion/exclusion criteria for each cohort may be found in Supplementary Information (Supplementary Tables S1-3, Supplementary Figure S1, and Supplementary Material). All sites obtained approval from the appropriate local institutional review boards and ethics committees, and all study participants provided written informed consent.

### Image acquisition and pre-processing

Structural T1-weighted brain MRI scans of each participant were acquired at each site. We used standardized protocols for image analysis and feature extraction (N_features_ = 153) across multiple cohorts (http://enigma.ini.usc.edu/protocols/imaging-protocols/). FreeSurfer [34] was used to segment and extract volumes bilaterally for 14 subcortical gray matter regions (nucleus accumbens, amygdala, caudate, hippocampus, pallidum, putamen, and thalamus), 2 lateral ventricles, along with 68 regional cortical thickness and 68 regional cortical surface area measures, and total intracranial volume (ICV). Cortical parcellations were based on the Desikan/Killiany atlas [35]. Segmentations were visually inspected and statistically examined for outliers. Further details of image acquisition parameters, software descriptions, and quality control may be found in Supplementary Table S4 and Supplementary Material.

### Brain age prediction

We used the publicly available ENIGMA brain age model (https://www.photonai.com/enigma_brainage). As described and discussed in Han *et al*. [24], brain age models were developed separately for males and females. The training samples were based on structural brain measures from 952 males and 1236 female healthy individuals (18-75 years of age) from the ENIGMA Major Depressive Disorder (MDD) group. There is no known participant overlap between the training samples and the participant data used in this work. Briefly, FreeSurfer measures from the left and right hemispheres were combined by calculating the mean ((left + right)/2)) of volumes for subcortical regions and lateral ventricles, and thickness and surface area for cortical regions, resulting in 77 features. The 77 average structural brain measures were used as predictors in a multivariable ridge regression to model chronological age in the healthy training samples (separately for males and females), using the Python-based *sklearn* package [36]. Model performance was originally validated in training samples (through 10-fold cross-validation) and out-of-sample controls. For more detailed information on the training samples, model development, and validation, see Supplementary Material and Han *et al*. [24]. Here, the parameters from the trained model were applied to our test samples of healthy controls and SZ patients to obtain brain-based age estimates for each cohort. To assess the model’s generalization performance in the test control samples, we calculated the (1) mean absolute error (MAE) between predicted brain age and chronological age, the (2) Pearson correlation coefficients between predicted brain age and chronological age (r), and (3) the proportion of chronological age variance explained by the model (R^2^).

### Statistical analyses

Brain-PAD (predicted brain-based age minus chronological age) was calculated for each participant and used as the outcome variable. While different prediction models were built for males and females, the generated brain-PAD values were pooled across sex for subsequent statistical analyses within each cohort. Each dependent measure of the i^th^ individual was modelled as follows

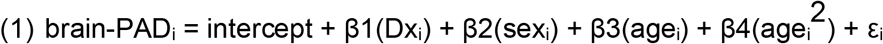

Where Dx represents diagnostic status for SZ. We corrected for the well-documented systematic age bias in brain age prediction (see Supplementary Material for brief explanation of this issue) [37, 38], as well as for potential confounding effects of age and sex in our test samples, by adding age, quadratic age (age^2^), and sex as covariates to our statistical models. We included both linear and quadratic age covariates in the same model as this provided a significantly better model fit to previous data compared with models including a linear age covariate only [24].

Within SZ patients, we also used linear models to examine associations between brain-PAD and clinical characteristics (CC):

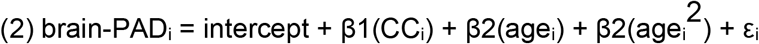

where “CC” represents either age of onset, illness duration (time from age-of-onset to time of scanning), SZ symptomatology at study inclusion (including Scale for the Assessment of Negative Symptoms – SANS Global, Scale for the Assessment of Positive Symptoms – SAPS Global, and Positive and Negative Syndrome Scale – PANSS Total), antipsychotic (AP) medication use at time of scanning (typical/atypical/both/none) or chlorpromazine (CPZ) dose equivalents (mg per day). All analyses were also repeated while covarying for handedness (right/left/ambidextrous). Cohorts with less than 5 healthy controls or SZ patients with respect to each clinical characteristic were excluded from the main or additional analyses (for exclusions, see Supplementary Material).

Cohort-specific results were then meta-analysed using the *rma* function in the *metafor* package [39]. Random (or mixed) effects models were fitted using restricted maximum likelihood estimation and inverse-variance weighting. Statistical tests were two-sided, and results for the effects of 9 clinical characteristics among SZ patients were false discovery rate (FDR) corrected (using the Benjamini-Hochberg procedure) and considered statistically significant at α < 0.05. In addition, as some cohorts were on average younger (or older) than others, or collected through multiple scanning sites (ASRB, FBIRN, Huilong, MCIC, MPRC, PAFIP) or different MRI scanners, post-hoc meta-regressions were performed to explore between-study heterogeneity in effect size with respect to the number of scanning sites (i.e., single vs. multi-site status), scanner field strength (i.e., 1.5T vs. 3T MRI), or mean sample age (across cases and controls).

Finally, to better understand the contribution or importance of individual structural brain measures for making brain age predictions, we calculated Pearson’s correlation coefficients between brain-predicted age and each of the 77 FreeSurfer features in each cohort. A weighted average by sample size across cohorts was then calculated for each correlation coefficient and plotted on cortical maps for illustrative purposes only. Correlation analyses were also conducted separately for SZ patients and healthy controls.

## Results

### Sample characteristics

Demographics and clinical characteristics across cohorts can be found in Table 1. Mean age weighted by sample size (range) across SZ patient and healthy control cohorts was 34.22 (18.36-43.66) and 33.82 (22.58-41.41) years, respectively. Patient and control cohorts were on average 67.32% (43.75-100) males and 54.89% (38.46-100) females, respectively. Weighted mean age of onset and duration of illness across patient cohorts were 24.75 (17.55-29.99) and 10.83 (0.62-18.87) years. Mean symptom severity (PANSS total) was 62.41 (33.38-93.12). For cohorts where current antipsychotic medication type information was available, the weighted mean percentage of patients on first-generation (typical), second-generation antipsychotics (atypical), both typical and atypical, or no antipsychotic medication was 10.05%, 67.65%, 14.73% and 7.57%, respectively.

**Table 1.**
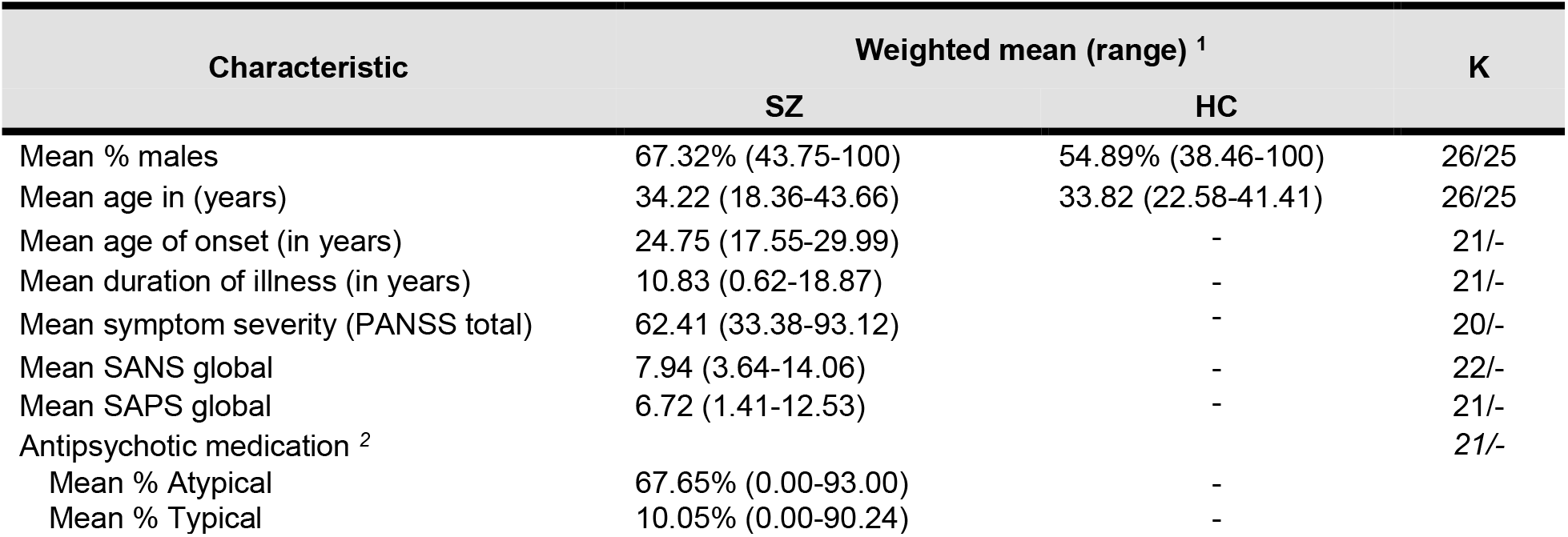
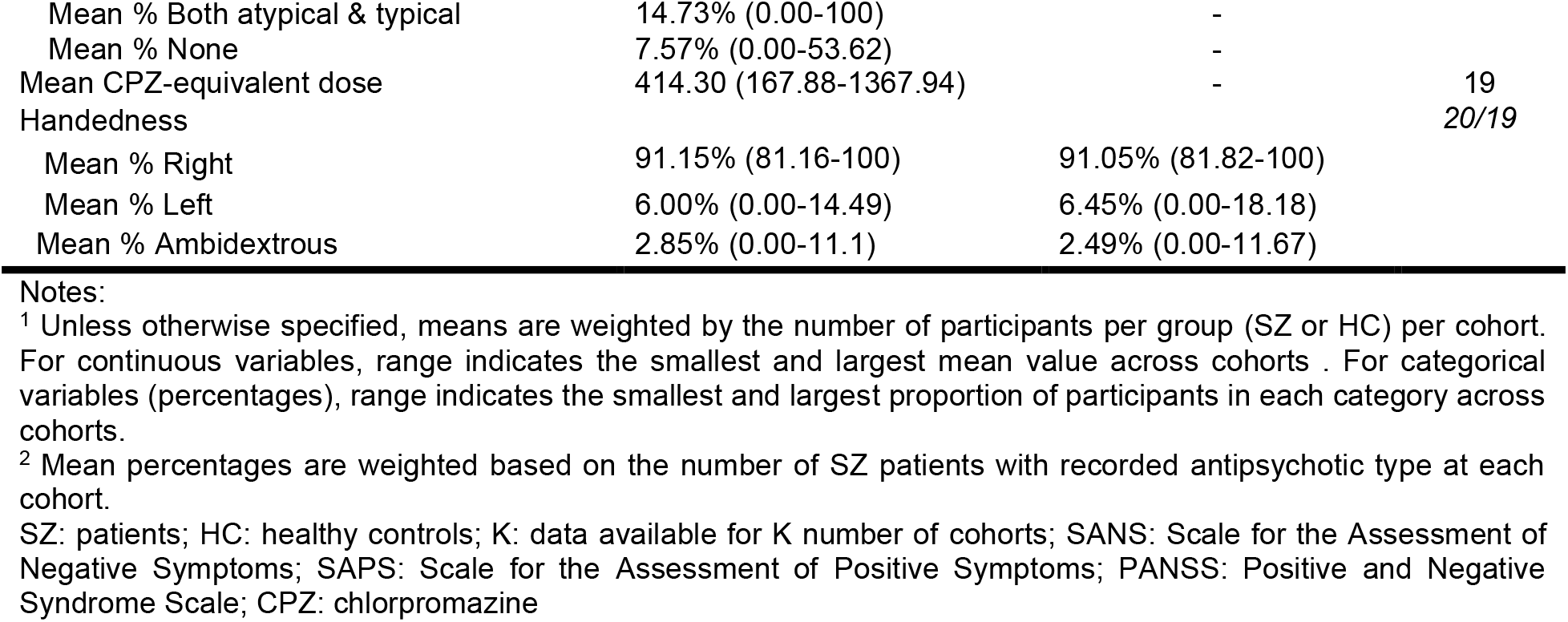
Participant characteristics for patients and controls across cohorts.

| Characteristic | Weighted mean (range) <sup>1</sup> |  | K |
| --- | --- | --- | --- |
|  | SZ | HC |  |
| Mean % males | 67.32% (43.75-100) | 54.89% (38.46-100) | 26/25 |
| Mean age in (years) | 34.22 (18.36-43.66) | 33.82 (22.58-41.41) | 26/25 |
| Mean age of onset (in years) | 24.75 (17.55-29.99) | - | 21/- |
| Mean duration of illness (in years) | 10.83 (0.62-18.87) | - | 21/- |
| Mean symptom severity (PANSS total) | 62.41 (33.38-93.12) | - | 20/- |
| Mean SANS global | 7.94 (3.64-14.06) | - | 22/- |
| Mean SAPS global | 6.72 (1.41-12.53) | - | 21/- |
| Antipsychotic medication <sup>2</sup> |  |  | 21/- |
| Mean % Atypical | 67.65% (0.00-93.00) | - |  |
| Mean % Typical | 10.05% (0.00-90.24) | - |  |
| Mean % Both atypical & typical | 14.73% (0.00-100) | - |  |
| Mean % None | 7.57% (0.00-53.62) | - |  |
| Mean CPZ-equivalent dose | 414.30 (167.88-1367.94) | - | 19 |
| Handedness |  |  | 20/19 |
| Mean % Right | 91.15% (81.16-100) | 91.05% (81.82-100) |  |
| Mean % Left | 6.00% (0.00-14.49) | 6.45% (0.00-18.18) |  |
| Mean % Ambidextrous | 2.85% (0.00-11.1) | 2.49% (0.00-11.67) |  |

### Brain age prediction performance

In controls, the weighted average MAE across cohorts was 7.60 (SE = ± 0.40) and 8.45 (SE = ± 0.46) years for males and females, respectively (Supplementary Figs. S2a-b). Within the SZ group, the MAE was 10.14 (SE = ± 0.52) and 9.61 (SE = ± 0.54) years for males and females, respectively (Supplementary Figs. S2c-d). Correlations between chronological age and predicted brain age were moderate to large in controls (males *r* = 0.64, and females *r* = 0.63; both R^2^ = 0.41), and in SZ patients (males *r* = 0.58, and females *r* = 0.62; both R^2^ = 0.33) (Supplementary Figs. S3a-d).

### Brain age differences between SZ and controls

Weighted mean brain-PAD scores were +4.39 years (SE= ±0.84) in the control group and +7.74 years (SE= ±0.94) in the SZ group. On average, brain-PAD was higher by +3.64 years (95% CI 3.01, 4.26; p <0.0001) in individuals with SZ compared to controls (Cohen’s d=0.50; 95% CI 0.34, 0.65; p<0.0001) adjusted for age, age^2^, and sex (Figure 1). Post-hoc sensitivity analysis excluding cohorts in which the model generalised less well (based on MAE > 10.00 or R^2^<0.1 in healthy controls) returned similar results (see Supplementary Fig. S4). Effect sizes were heterogeneous across individual cohorts (Q (24) = 52.52, p<0.0007; I^2^ =55.28%). A significant effect was seen in 22 out of 25 cohorts, with a positive direction of mean effect size observed in all but one cohort. Across cohorts, mean brain-PAD did not differ between single versus multi-site cohorts (QM(1)=0.061, p=0.805), nor between 1.5T versus 3T scanners (QM(1)=0.053; p=0.818) or with respect to mean age (QM(1)=0.248, p= 0.619). We also found a weak linear, yet not significant effect for age on brain-PAD scores (b_age_ = -0.24, 95% CI -0.48, 0.00, p=0.054; b_age2_ = -0.00, 95%CI -0.03, 0.03, p=0.855). Additional adjustment for handedness in a smaller pool of 16 cohorts did not meaningfully change our main finding for the effect of SZ (+3.64 years; 95% CI 2.84, 4.44; p<0.0001).

**Figure 1.**
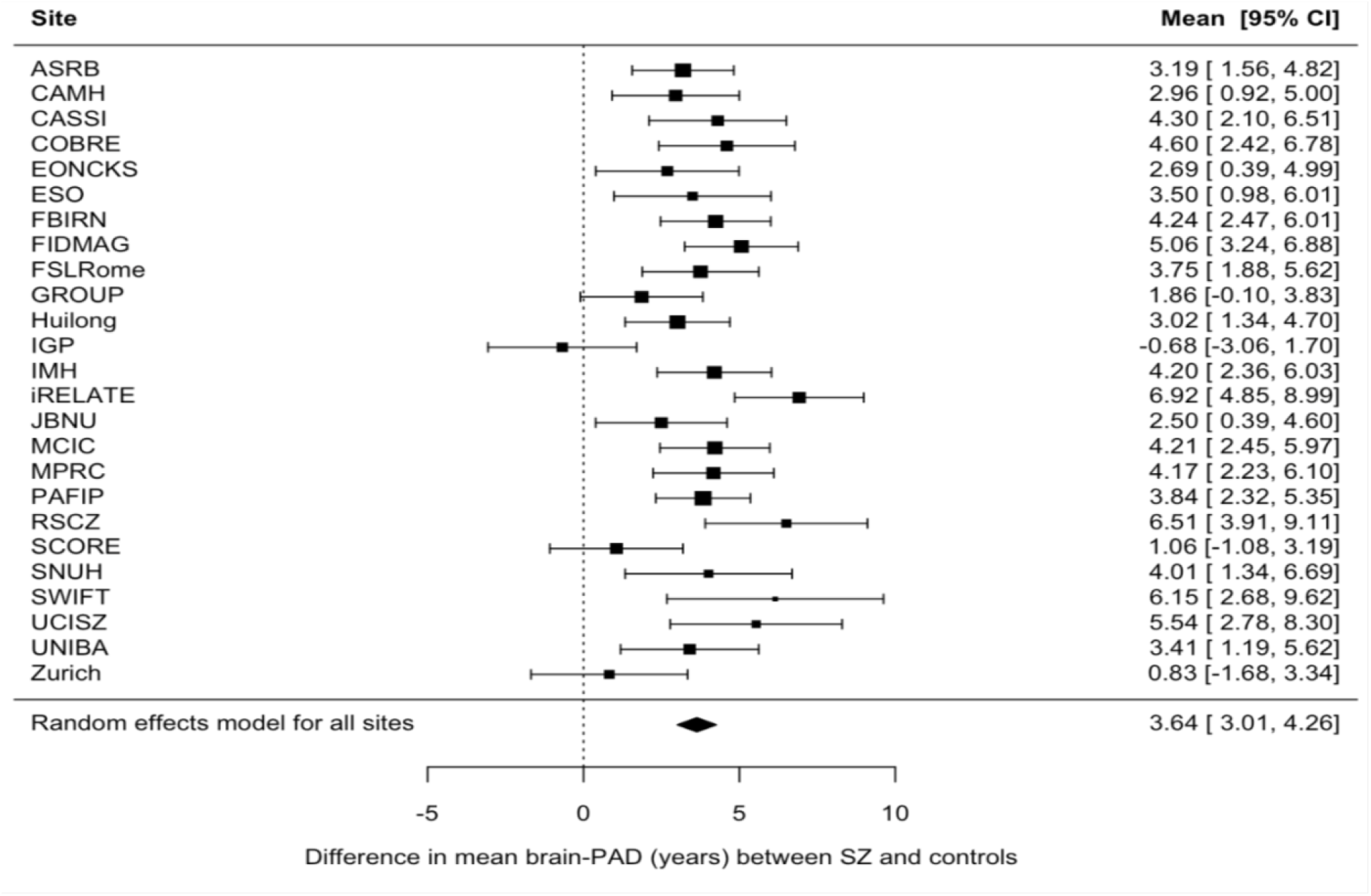
Case-control differences in brain-PAD. Forest plot of differences in mean brain-PAD scores (predicted brain age - chronological age) between patients with schizophrenia (SZ) and controls across (26 -1) 25 cohorts (a total of 2792 cases and 2598 controls; excluding 1 cohort that contributed data for patients only), controlling for sex, age and age^2^. Regression coefficients (in years) are denoted by black boxes. Black lines indicate 95% confidence intervals. The size of the box indicates the weight the cohort received (based on inverse variance weighting). The pooled estimate for all cohorts is represented by a black diamond, with the outer edges of the diamond indicating the confidence interval limits.

### Brain age and clinical characteristics in SZ

Among SZ patients, we found no statistically significant effects on brain-PAD of clinical characteristics, including age-of-onset, length of illness, symptom severity (PANSS total, SAPS global), antipsychotic use, and CPZ-equivalent dose after adjusting for age and age^2^ (Table 2 and Supplementary Figs. S5a-i). A weak, positive effect for negative symptom severity (SANS global) on Brain-PAD was observed, although it did not reach significance (b=0.18, 95% CI -0.01, 0.38, PFDR=0.62). In addition, no significant effects were found for typical versus atypical and both atypical and typical versus atypical medication groups (Supplementary Table S5). Further adjustment for handedness returned similar results (Supplementary Table S6).

**Table 2.** Clinical characteristics and brain-PAD in individuals with SZ

| Clinical parameter | N | K | beta | SE | 95% CI | P<sub>FDR</sub> value |
| --- | --- | --- | --- | --- | --- | --- |
| Age of onset (years) | 2053 | 21 | -0.06 | 0.09 | -0.22, 0.11 | 0.84 |
| Length of illness (years) | 2056 | 21 | 0.05 | 0.09 | -0.12, 0.22 | 0.84 |
| PANSS total | 1437 | 20 | 0.05 | 0.06 | -0.06, 0.17 | 0.77 |
| SANS global | 1911 | 22 | 0.18 | 0.10 | -0.01, 0.38 | 0.62 |
| SAPS global | 1892 | 21 | 0.14 | 0.12 | -0.09, 0.38 | 0.70 |
| AP use – atypical vs. unmed | 642 (486/156) | 7 | 1.71 | 1.27 | -0.77, 4.19 | 0.70 |
| AP use – typical vs. unmed | 117 (42/72) | 3 | -0.13 | 1.00 | -2.10, 1.84 | 0.90 |
| AP use – both vs. unmed | 266 (184/82) | 4 | -0.33 | 1.08 | -2.43, 1.77 | 0.90 |
| CPZ-equivalent dose | 1698 | 19 | 0.00 | 0.01 | -0.02, 0.02 | 0.90 |

Associations between clinical characteristics and brain-PAD (predicted brain age – chronological age) in SZ. For continuous variables (age of onset, length of illness, PANSS total and CPZ), the regression coefficient *beta* indicates a mean change in brain-PAD per unit increase in each clinical variable across cohorts. For categorical variables (AP use – typical/atypical/both atypical and typical), *beta* indicates the mean brain-PAD difference between each treatment group and unmedicated (unmed) individuals. Effects were adjusted for age and age<sup>2</sup>. K: number of cohorts; N: total number of participants included in each meta-analysis (where applicable, total group size for AP type use/unmedicated is given in brackets); SE: standard error; CI: confidence intervals. P values are false discovery rate (FDR) adjusted. SANS: Scale for the Assessment of Negative Symptoms; SAPS: Scale for the Assessment of Positive Symptoms; PANSS: Positive and Negative Syndrome Scale; AP: Antipsychotics; CPZ: chlorpromazine.

### Correlations between brain imaging features and brain age

All imaging features, except mean lateral ventricle volume, were negatively correlated with predicted brain age (Figure 2); thickness features correlated more strongly with brain age (mean Pearson r [SD]: −0.46 [0.13]), especially in medial frontal and temporo-parietal regions, than subcortical volumes (−0.32 [0.30]) or surface area features (−0.22 [0.06]). We also visualized these associations separately for controls and SZ patients with similar results, suggesting comparable structure coefficients in both groups (for more details see Supplementary Material).

**Figure 2.**
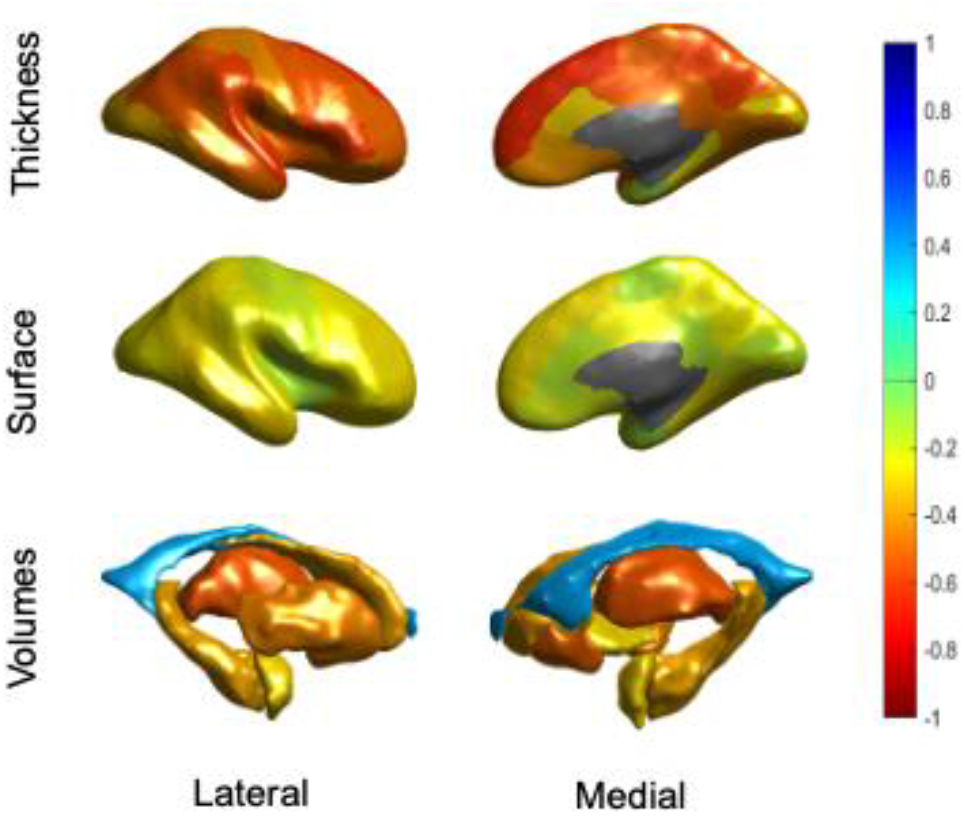
Correlation coefficients of predicted brain age and FreeSurfer features across control and schizophrenia (SZ) groups. Bivariate correlations are shown to provide an indication of the relative contribution of features in brain age prediction. The figure shows Pearson correlations between predicted brain age and cortical thickness features (top row), cortical surface areas (middle row) and subcortical volumes (bottom row), from both the lateral (left) and medial (right) view. Features were averaged across the left and right hemispheres. The negative correlation with ICV was excluded from this figure for display purposes.

## Discussion

We assessed brain ageing in 2803 individuals with SZ and 2598 healthy controls using a novel brain age algorithm based on FreeSurfer ROIs. Results indicate that, at a group level, patients with SZ show a greater discrepancy between their brain-predicted age and chronological age compared to healthy individuals (+3.64 years), with a moderate increase in brain-PAD (Cohen’s=0.50). The greater brain-PAD in the SZ group was not driven by any of the specific clinical characteristics assessed here (age of onset, length of illness, symptom severity, and antipsychotic use and dose). This study has two major strengths. Firstly, through a prospective meta-analytic approach within the ENIGMA consortium, we were able to assess brain age differences between SZ patients and healthy controls using standardised analysis methods across multiple independent cohorts worldwide, providing a generalised mean effect size. Second, the overall large sample size and harmonisation of data across cohorts allowed for a more reliable assessment of the relationship between clinical variables and brain-PAD among SZ patients.

The mean brain-PAD difference between patients and controls was +3.64 years (Cohen’s d=0.50) in our study. Overall, this finding is aligned with previously reported brain-PAD scores in SZ patients vs. healthy controls (range: +2.6-7.8 years) [26–33]. Schnack *et al*. [29] and a recent mega-analysis by Kaufmann *et al*. [32] found similar effect sizes (+3.4 years and Cohen’s d = 0.51, respectively) in largely non-overlapping/independent samples from this current study. On the other hand, our brain-PAD difference is smaller relative to that reported in earlier work by Koutsouleris *et al*. [27] and Shahab *et al*. [30] showing respectively +5.5 to +7.8 years of brain age in smaller samples of SZ patients. Several methodological differences may explain the variability in magnitude of brain age effects in SZ across studies, including the type of neuroimaging features (e.g., voxel-wise vs. ROI-based morphometric data; and/or single vs. multiple imaging modalities) [40], the machine learning algorithm used for brain age estimation [41], the size of training and test data samples, and differences in patient characteristics.

Relative to healthy controls, brain-PAD scores in SZ suggest more advanced brain ageing than in MDD (+1.12 years) [42] and bipolar disorder (BP; +1.93 years) [42], that may reflect more pronounced structural brain abnormalities in SZ [24]. This aligns with previous reports from the ENIGMA consortium, showing largest effect sizes of cortical and subcortical gray matter alterations in SZ (highest Cohen’s d effect size=0.53) [16, 17], followed by BD (highest Cohen’s d=0.32) [43, 44] and MDD (highest Cohen’s d=0.14) [45, 46]. Hence, sensitivity of brain-PAD to SZ at the group level appears to be quantitively similar to that of leading cortical thickness and subcortical volume measures. A further key advantage of the “brain age” paradigm is that it captures multivariate age-related structural brain patterns into one (or more) composite measure(s), thereby simplifying analyses and aids interpretation with respect to normative patterns of brain ageing.

Consistent with previous reports [27, 31], we did not observe significant associations between brain-PAD and age of onset, length of illness, and antipsychotic treatment or dose among SZ patients. This suggests that a greater brain-PAD in SZ may not be primarily driven by disease progression or treatment-related effects on brain structure that have been reported elsewhere [12, 14, 18, 47, 48]. This is in keeping with previous studies showing a greater brain-PAD already present in first-episode SZ and first-episode psychosis patients [26, 49]. Using a longitudinal design, Schnack *et al*. investigated brain age acceleration (i.e., annual rate of change in brain-PAD) over the duration of illness in SZ (N=341; mean follow up period: 3.48 years). Brain-PAD started increasing by about 2.5 years (per year) just after illness onset, though this acceleration rate slowed down to a normal rate over the first 5 years of illness [29]. Lastly, in contrast to previous findings in SZ [27] and first-episode psychosis [49] we did not find strong evidence for a positive association between negative symptom severity and brain-PAD. An explanation for this could be that negative symptoms are more specifically linked to brain age differences at the regional level (i.e., temporal or parietal brain-PAD) than at the global level (i.e., ‘whole-brain’ brain-PAD), as reported previously [32].

The biological mechanisms underlying advanced brain ageing in SZ remain elusive. These may involve interrelated biochemical abnormalities that accompany both schizophrenia and brain ageing, including increased inflammation and oxidative stress [10, 50]. Elevated levels of inflammatory markers (e.g., pro-inflammatory cytokines in blood and central nervous system) have been observed by multiple studies in individuals with schizophrenia [11, 51]. Moreover, there has been evidence for peripheral inflammation markers being associated with structural brain abnormalities in schizophrenia and related outcomes (e.g., first episode psychosis), including but not limited to abnormal cortical thickness of the bilateral Broca’s area and temporal gyrus [52, 53], as well as with greater brain-PAD scores [54]. Abnormal levels of multiple oxidative stress markers have also been observed in SZ, both peripherally and in brain tissue [11, 55]. Oxidative stress and inflammation may reciprocally induce one another via a positive feedback loop in SZ, resulting in cellular damage [56].

Several methodological issues require further consideration. First, while a brain-PAD score (that is not equal to zero) is conceptually a prediction error that could reflect physiological deviations from normal ageing trajectories, it could be partly attributed to lack of model accuracy due to noise or unwanted variation [32, 57, 58]. Potential sources of unwanted variation include the use of multiple scanners and/or image acquisition protocols across (or within) participating cohorts that may affect the overall generalization performance of the brain age model applied here. Nevertheless, while our model fit is lower than some previous studies, this would only increase noise, not a bias towards finding an effect of SZ on brain-PAD, as each cohort included in the primarily analysis had data on both cases and healthy controls that were collected in a similar, if not identical, manner (e.g., same site/scanner and/or image acquisition protocol). Second, although our meta-analytic approach allowed us to combine information across multiple cohorts, the summary-level data reported here does not adequately capture the considerable inter-individual variability in brain-PAD among SZ patients, as has been documented elsewhere [32]. As some individuals with SZ are not characterised by a greater brain-PAD, it would be important to further investigate both clinical as well as biological and lifestyle factors that are linked to SZ (e.g., inflammation, smoking, body mass index) potentially accounting for inter-individual variability. Given that greater brain-PAD has been associated with poorer health outcomes, such as an increased mortality risk [23], understanding the extent to which various factors may contribute to brain ageing in SZ could help prioritize targets for interventions aiming to halt (or reverse) advanced brain ageing. Third, although the sample size of our main analysis (SZ versus controls) was very large for a neuroimaging study, the size of patient groups categorised by status of antipsychotic use was relatively small (particularly that of unmedicated individuals with SZ) and sample differences include the use of different assessments or processes to ascertain medication use and dose. This may have precluded detection of some associations. Lastly, given the cross-sectional design of the current study, we were not able to assess brain age acceleration more directly and how that may be related to clinical characteristics. Longitudinal large-scale studies are better suited for examining brain ageing *per se* [59] and for evaluating the clinical relevance of brain-PAD in SZ.

In conclusion, we found evidence of advanced brain ageing in SZ patients compared to healthy controls, which does not seem to be driven by the effects of medication or other clinical characteristics. Deviations from normative brain ageing trajectories in SZ may at least in part reflect increased risk of premature mortality and age-related chronic diseases commonly seen in SZ. Future longitudinal studies with more in-depth clinical characterization - including information on mental and somatic health outcomes - will be needed to elucidate whether a brain age predictor such as brain-PAD can provide a clinically useful biomarker to inform early prevention or intervention strategies in SZ.

## Supporting information

STROBE Checklist

## Data Availability

All data produced in the present study are available upon reasonable request to the authors.

## Acknowledgments

The ENIGMA Schizophrenia Working Group acknowledges the National Institute of Health (NIH) Big Data to Knowledge (BD2K) award for foundational support and consortium development (Grant No. U54 EB020403 to PMT). For a complete list of ENIGMA-related grant support please see here: http://enigma.ini.usc.edu/about-2/funding/. The content of this manuscript is solely the responsibility of the authors and does not necessarily represent the official views of the National Institutes of Health. CC was supported by grant MR/N0137941/1 for the GW4 BIOMED Doctoral Training Partnership awarded to the Universities of Bath, Bristol, Cardiff and Exeter from the Medical Research Council (MRC)/UKRI. JC was supported by a UK Research & Innovation (UKRI) Innovation Fellowship (MR/R024790/2). TH was funded by the German Research Foundation (DFG grants HA7070/2-2, HA7070/3, HA7070/4 to TH). NJ and LS were supported by NIH grant R01 MH117601. NJ was also supported by NIH grants R01AG059874, R01MH11760. LS was also supported by an NHMRC Career Development Fellowship (1140764) and a University of Melbourne Dame Kate Campbell fellowship. PMT was supported in part by NIH grant R01 MH116147. EW was supported by the European Union Horizon 2020 research and innovation programme (EarlyCause, grant number 848158). Acknowledgements for the various participating data contributors follow - ASRB: the Australian Schizophrenia Research Bank (ASRB) was supported by the National Health and Medical Research Council of Australia (NHMRC) (Enabling Grant, ID 386500), the Pratt Foundation, Ramsay Health Care, the Viertel Charitable Foundation and the Schizophrenia Research Institute. Chief Investigators for ASRB were Carr, V., Schall, U., Scott, R., Jablensky, A., Mowry, B., Michie, P., Catts, S., Henskens, F., Pantelis, C. We thank Loughland, C., the ASRB Manager, and acknowledge the help of Jason Bridge for ASRB database queries. CP was also supported by a National Health and Medical Research Council (NHMRC) Senior Principal Research Fellowship (1105825), an NHMRC L3 Investigator Grant (1196508). CSW was funded by the NSW Ministry of Health, Office of Health and Medical Research and a recipient of a National Health and Medical Research Council (Australia) Principal Research Fellowship (PRF) (#1117079). CAMH: the datasets were generated and shared with support from the CAMH Foundation and the Canadian Institutes of Health Research. CASSI: this study was supported by NHMRC grant 568807 (awarded to CSW and TW). CSW was funded by the NSW Ministry of Health, Office of Health and Medical Research and a recipient of a National Health and Medical Research Council (Australia) Principal Research Fellowship (PRF) (#1117079). COBRE: the study and investigators were supported by NIH grants R01EB006841 &P20GM103472, as well as NSF grant 1539067. JT (senior author) was supported by NIH grant R01MH121246. EONCKS: all processing of contributed neuroimaging data was conducted on the Centre for High Performance Computing, Cape Town, South Africa. ESO: the study and investigators (AC, FS, DT) was funded by the Ministry of Health, Czech Republic - Conceptual Development of Research Organization (Institute for Clinical and Experimental Medicine – IKEM, IN 00023001), and Ministry of Health of the Czech Republic, grant nr. NU20-04-00393. FBIRN: the study was supported by the National Center for Research Resources at the National Institutes of Health (NIH 1 U24 RR021992 (Function Biomedical Informatics Research Network) and NIH 1 U24 RR025736-01 (Biomedical Informatics Research Network Coordinating Center; http://www.birncommunity.org). FBIRN data was processed by the UCI High Performance Computing cluster supported by Joseph Farran, Harry Mangalam, and Adam Brenner and the National Center for Research Resources and the National Center for Advancing Translational Sciences, National Institutes of Health, through Grant UL1 TR000153. JT (senior author) was also supported by NIH grant R01MH121246. FIDMAG: this was supported by the CIBERSAM and the Catalonian Government, Generalitat de Catalunya (2017-SGR-1271 to EP-C from AGAUR). PFC was supported by a Sara Borrell contract (CD19/00149), funded by Instituto de Salud Carlos III, co-funded by the European Union (European Regional Development Fund/European Social Fund [ERDF/ESF], “Investing in your future”). FSLRome: the IRCCS Santa Lucia Foundation of Rome study (FSLRome) was partially supported by the Italian Ministry of Health (RC12-13-14-15A Grant) and by the European Commission ERA-Net NEURON joint transnational calI 2010 (European Research Projects on Mental Disorders: NEUCONNECT). GROUP: This work was supported by the Geestkracht program of the Dutch Health Research Council (ZON-MW, grant number: 10-000-1002), and the European Community’s Seventh Framework Program under grant agreement No. HEALTH-F2-2009-241909 (Project EU-GEI). Huilong: this study was funded by the National Natural Science Foundation of China (81761128021;31671145;81401115;81401133), Beijing Municipal Science & Technology Commission grant (Z141107002514016) and Beijing Natural Science Foundation (7162087, Beijing Municipal Administration of Hospitals Clinical medicine Development of special funding (XMLX201609; zylx201409). IGP: this study was supported by National Health and Medical Research Council (NHMRC) Project Grant APP630471, APP1051673, APP1081603 (awarded to MG). IMH: this study was supported by the National Healthcare Group, Singapore (SIG/05004; SIG/05028) and the Singapore Bioimaging Consortium (RP C009/06) research grants (awarded to KS). iRELATE: this study was supported by grants to GD from the European Research Council (ERC-2015-STG-677467) and Science Foundation Ireland (SFI-16/ERCS/3787). Madrid: this research was supported by the Spanish Ministry of Science and Innovation; Instituto de Salud Carlos III (grants SAM16PE07CP1 and PI16/02012 awarded to CAra; PI17/01249 awarded JJ; and PI17/00481, PI19/00024 and PI20/00721 awarded to CMD-C), co-financed by ERDF Funds from the European Commission, “A way of making Europe”, CIBERSAM; Madrid Regional Government (B2017/BMD-3740 AGES-CM-2 awarded to CAra), European Union Structural Funds; European Union Seventh Framework Program under grant agreements FP7-4-HEALTH-2009-2.2.1-2-241909 (Project EU-GEI), FP7-HEALTH-2013-2.2.1-2-603196 (Project PSYSCAN) and FP7-HEALTH-2013-2.2.1-2-602478 (Project METSY) awarded to Cara; and European Union H2020 Program under the Innovative Medicines Initiative 2 Joint Undertaking (grant agreement No 115916, Project PRISM, and grant agreement No 777394, Project AIMS-2-TRIALS awarded to CAra), Fundación Familia Alonso and Fundación Alicia Koplowitz. MCIC: the study was supported by the National Institutes of Health (NIH/NCRR P41RR14075 and R01EB005846 (to Vince D. Calhoun)), the Department of Energy (DE-FG02-99ER62764), the Mind Research Network, the Morphometry BIRN (1U24, RR021382A), the Function BIRN (U24RR021992-01, NIH.NCRR MO1 RR025758-01, NIMH 1RC1MH089257 to Vince D. Calhoun). MPRC: support was received from NIH grants R01MH123163, R01EB015611, R01NS114628 (awarded to PK). PAFIP: this study has been funded by Instituto de Salud Carlos III through the project PI17/01056 (co-funded by European Regional Development Fund/European Social Fund “A way to make Europe”/”Investing in your future”). RSCZ: data collection was supported by RFBR 15-06-05758 grant. SCORE: this study was supported in part by grant 3232BO_119382 from the Swiss National Science Foundation. We thank the FePsy (Frueherkennung von Psychosen; early detection of psychosis) Study Group from the University of Basel, Department of Psychiatry, Switzerland, for the recruitment of the study participants. The FePsy study was supported in part by grant No. SNF 3200-057216/1, ext./2, ext./3. SNUH: this research was supported by the Basic Science Research Program and the Brain Research Program through the National Research Foundation of Korea (NRF) funded by the Ministry of Science & ICT (Grant no. 2019R1C1C1002457 and 2020M3E5D9079910 (awarded to MK and JSK). UNIBA: this study was supported by grant funding from the Italian Ministry of Health (PE-2011-02347951). PS was supported by a PhD studentship jointly funded by the NIHR-BRC at SLaM and the Department of Neuroimaging, King’s College London. Zurich: this study was supported by the Swiss National Science Foundation (Grant No. 105314_140351 and 10001CL_169783 to SK). MK acknowledges funding from the Swiss National Science Foundation (P2SKP3_178175).

