## Supplementary Tables and Figures for "Brain ageing in schizophrenia: evidence from 26 international cohorts via the ENIGMA Schizophrenia consortium"

### Contents

**Supplementary Table S1.** ENIGMA Schizophrenia Working Group - Sample demographics by cohort

**Supplementary Table S2.** ENIGMA Schizophrenia Working Group - Sample current medication information by cohort

**Supplementary Table S3.** ENIGMA Schizophrenia Working Group - Diagnostic Instrument and exclusion criteria by cohort

**Supplementary Table S4.** ENIGMA Schizophrenia Working Group - Image acquisition and processing by cohort

**Supplementary Table S5.** Additional results for antipsychotic use and brain-PAD in SZ

**Supplementary Table S6.** Clinical characteristics and brain-PAD in SZ, additionally adjusting for handedness

**Supplementary Figure S1.** ENIGMA Schizophrenia Working Group - World map showing the location of each sample included in this study

**Supplementary Figure S2a.** Mean absolute error for healthy control males across cohorts

**Supplementary Figure S2b.** Mean absolute error for healthy control females across cohorts

**Supplementary Figure S2c.** Mean absolute error for schizophrenia males across cohorts

**Supplementary Figure S2d.** Mean absolute error for schizophrenia females across cohorts

**Supplementary Figure S3a.** Correlation between brain-predicted age and chronological age in healthy control males across cohorts

**Supplementary Figure S3b.** Correlation between brain-predicted age and chronological age in healthy control females across cohorts

**Supplementary Figure S3c.** Correlation between brain-predicted age and chronological age in schizophrenia males across cohorts

**Supplementary Figure S3d.** Correlation between brain-predicted age and chronological age in schizophrenia females across cohorts

**Supplementary Figure S4.** Sensitivity analysis of case-control differences in brain-PAD

**Supplementary Figure S5a.** Age of illness onset and brain-PAD across cohorts

**Supplementary Figure S5b.** Duration of illness and brain-PAD across cohorts

**Supplementary Figure S5c.** Overall illness severity (PANSS total) and brain-PAD across cohorts

**Supplementary Figure S5d.** Negative symptom severity (SANS global) and brain-PAD across cohorts

**Supplementary Figure S5e.** Positive symptom severity (SAPS global) and brain-PAD across cohorts

**Supplementary Figure S5f.** Current atypical antipsychotic use and brain-PAD across cohorts

**Supplementary Figure S5g.** Current typical antipsychotic use and brain-PAD across cohorts

**Supplementary Figure S5h.** Current (both) typical & atypical antipsychotic use and brain-PAD across cohorts

**Supplementary Figure S5i.** Current antipsychotic daily dose and brain-PAD across cohorts

**Supplementary Table S1**. ENIGMA Schizophrenia Working Group - Sample Demographics by participating cohort

| Cohort | N (T) | N (SZ) | N (HC) | M/F (SZ) | M/F (HC) | Mean Age (SZ) | | Mean Age (HC) | | Mean Age of Onset (years) | | Mean Duration Of Illness (years) | | PANSS Total | | SANS Global | | SAPS Global | | Handedness (R/L/A) (SZ) | Handedness (R/L/A) (HC) |
|---|---|---|---|---|---|---|---|---|---|---|---|---|---|---|---|---|---|---|---|---|---|
| ASRB | 427 | 262 | 165 | 176/86 | 78/87 | 38.63 | ± 10.32 | 39.36 | ± 13.60 | 23.60 | ± 6.59 | 15.03 | ± 9.57 | - | - | - | - | - | - | 239/23/0 | 135/30/0 |
| CAMH | 253 | 116 | 137 | 69/47 | 72/65 | 43.39 | ± 16.09 | 41.20 | ± 17.09 | 24.63 | ± 8.50 | 18.87 | ± 15.7 | 53.47 | ±15.44 | 7.31 | ±3.90 | 4.73 | ±3.26 | 105/8/3 | 131/6/0 |
| CASSI | 116 | 53 | 63 | 35/18 | 33/30 | 35.17 | ± 8.75 | 30.49 | ± 7.08 | 23.08 | ± 5.69 | 12.11 | ± 7.12 | 33.38 | ±16.43 | 3.64 | ±3.98 | 1.41 | ±2.88 | 45/4/3 | 53/4/4 |
| COBRE | 135 | 69 | 66 | 56/13 | 46/20 | 37.41 | ± 13.38 | 36.11 | ± 11.93 | 21.84 | ± 8.09 | 15.34 | ±12.47 | 59.68 | ±15.19 | 7.75 | ±2.97 | 5.38 | ±2.91 | 56/10/3 | 63/1/2 |
| EONCKS | 176 | 90 | 86 | 66/24 | 52/34 | 24.96 | ± 6.64 | 26.95 | ± 7.21 | - | - | - | - | 93.12 | ±15.24 | 14.06 | ±4.10 | 10.62 | ±2.54 | - | - |
| ESO | 69 | 34 | 35 | 17/17 | 17/18 | 29.68 | ± 7.33 | 28.74 | ± 6.47 | 28.97 | ± 7.22 | 0.62 | ± 0.82 | 63.41 | ±18.32 | 8.53 | ±3.46 | 4.75 | ±3.43 | - | - |
| FBIRN | 357 | 183 | 174 | 137/46 | 124/50 | 38.83 | ± 11.6 | 37.52 | ± 11.26 | 21.72 | ± 7.65 | 17.17 | ± 11.58 | 58.64 | ±15.07 | 7.61 | ±3.77 | 5.55 | ±2.90 | 166/13/4 | 165/7/2 |
| FIDMAG | 283 | 160 | 123 | 124/36 | 54/69 | 39.64 | ± 11.86 | 37.54 | ± 10.13 | 23.01 | ± 6.99 | 15.53 | ± 11.35 | 76.21 | ±18.19 | 12.95 | ±4.50 | 6.32 | ±3.24 | 160/0/0 | 123/0/0 |
| FSLRome | 278 | 162 | 116 | 109/53 | 73/43 | 39.68 | ± 11.10 | 37.48 | ± 11.52 | 24.61 | ± 8.50 | 15.09 | ± 10.63 | 86.07 | ±20.75 | 10.42 | ±5.00 | 7.99 | ±3.71 | 151/5/2 | 87/7/3 |
| GROUP | 260 | 85 | 175 | 58/27 | 79/96 | 28.36 | ± 6.84 | 30.68 | ± 9.56 | - | - | - | - | - | - | - | - | - | - | - | - |
| Huilong | 320 | 234 | 86 | 128/106 | 47/39 | 25.94 | ± 5.76 | 27.97 | ± 6.61 | - | - | - | - | - | - | - | - | - | - | - | - |
| IGP | 107 | 46 | 61 | 27/19 | 31/30 | 41.50 | ± 11.09 | 35.22 | ± 10.95 | 23.13 | ± 7.62 | 17.97 | ± 9.94 | 56.93 | ±18.59 | 8.02 | ±4.6 | 4.42 | ±3.38 | 39/1/4 | 52/1/7 |
| IMH | 227 | 151 | 76 | 105/46 | 47/29 | 33.08 | ± 9.11 | 31.75 | ± 9.80 | 25.92 | ± 7.49 | 7.53 | ± 7.04 | 39.91 | ±8.34 | 3.91 | ±2.03 | 2.80 | ±2.17 | 138/12/1 | 68/8/0 |
| iRELATE | 215 | 50 | 165 | 36/14 | 96/69 | 43.66 | ± 10.81 | 36.33 | ± 12.64 | 27.74 | 8.98 | 17.57 | 10.71 | 38.52 | ±8.38 | 4.43 | ±2.6 | 1.61 | ±1.24 | 45/3/2 | 135/16/1 |
| JBNU | 208 | 94 | 114 | 57/37 | 48/66 | 39.29 | ± 10.22 | 41.41 | ± 9.86 | 29.99 | ± 9.08 | 9.03 | ± 7.88 | 52.96 | ±14.88 | 6.33 | ±3.54 | 4.85 | ±3.52 | 88/2/4 | 113/0/0 |
| Madrid | 11 | 11 | 0 | 10/1 | 0/0 | 18.36 | ± 0.50 | - | - | 17.55 | ± 0.82 | 0.82 | ± 0.75 | 89.18 | ±36.41 | 13.83 | ±6.88 | 8.89 | ±7.43 | 9/0/0 | - |
| MCIC | 311 | 148 | 163 | 113/35 | 101/62 | 32.89 | ± 10.51 | 31.43 | ± 11.05 | 22.75 | ± 6.60 | 10.22 | ± 9.75 | - | - | 7.95 | ±4.1 | 6.76 | ±3.16 | 131/7/6 | 149/5/9 |
| MPRC | 413 | 199 | 214 | 126/73 | 93/121 | 36.13 | ± 12.48 | 38.29 | ± 14.05 | - | - | - | - | - | - | - | - | - | - | - | - |
| PAFIP | 540 | 343 | 197 | 208/135 | 121/76 | 30.18 | ± 8.66 | 29.66 | ± 7.38 | 29.21 | ± 8.63 | 1.02 | ± 1.94 | 63.52 | ±16.87 | 5.81 | ±5.55 | 12.53 | ±3.43 | 286/22/21 | 175/12/9 |
| RSCZ | 88 | 40 | 48 | 40/0 | 48/0 | 22.83 | ± 2.86 | 22.75 | ± 2.53 | 21.71 | ± 2.69 | 1.12 | ± 1.32 | 58.05 | ±10.21 | 9.80 | ±2.76 | 2.93 | ±1.50 | 40/0/0 | 48/0/0 |
| SCORE | 121 | 69 | 52 | 49/20 | 23/29 | 26.87 | ± 6.73 | 26.08 | ± 3.89 | - | - | - | - | - | - | 8.83 | ±7.34 | - | - | - | - |
| SNUH | 72 | 32 | 40 | 14/18 | 20/20 | 24.44 | ± 5.22 | 22.58 | ± 3.94 | 23.81 | ± 5.25 | 6.70 | ± 4.03 | 67.00 | ±13.57 | 9.07 | ±3.30 | 6.10 | ±3.07 | 28/4/0 | 35/5/0 |
| SWIFT | 37 | 24 | 13 | 17/7 | 5/8 | 34.21 | ± 10.91 | 29.31 | ± 3.99 | 24.77 | ± 7.48 | 9.51 | ± 7.84 | 57.96 | ±12.05 | 6.41 | ±3.32 | 6.06 | ±3.04 | 24/0/0 | 13/0/0 |
| UCISZ | 57 | 27 | 30 | 22/5 | 23/7 | 42.93 | ± 10.62 | 41.37 | ± 12.26 | 25.00 | ± 7.76 | 17.5 | ± 10.05 | 59.96 | ±12.00 | 8.60 | ±3.91 | 5.60 | ±2.36 | 22/2/3 | 25/4/1 |
| UNIBA | 232 | 61 | 171 | 43/18 | 77/94 | 32.13 | ± 10.00 | 26.40 | ± 7.31 | 19.84 | ± 4.59 | 3.95 | ± 4.26 | 73.70 | ±22.67 | 8.92 | ±6.56 | 5.88 | ±3.27 | 52/1/2 | 138/12/9 |
| Zurich | 88 | 60 | 28 | 45/15 | 18/10 | 30.53 | ± 8.48 | 32.54 | ± 9.32 | 22.23 | ± 5.41 | 8.36 | ± 7.26 | 48.65 | ±10.41 | 7.58 | ±3.90 | 2.85 | ±1.51 | 53/7/0 | 24/4/0 |

N=Number; SZ=Schizophrenia; HC=Healthy Control; T=Total; M=Male; F=Female; PANSS=Positive and Negative Syndrome Scale; SANS=Schedule for the Assessment of Negative Symptoms; SAPS=Schedule for the Assessment of Positive Symptoms; R=right-handed; L=Left-handed; A= Ambidextrous; “-“: data not available for this cohort. Means are accompanied by standard deviation (±). Numbers for handedness per site may not add up to total sample and/or group (SZ/HC) size due to missing data

**Supplementary Table S2.** Current medication information (at the time of brain scanning) **by cohort**

| Cohort | N (unmedicated) | N Second Generation (atypical) | N First Generation (typical) | N Both (typical + atypical) | N NA | Mean CPZ | |
|---|---|---|---|---|---|---|---|
| ASRB | 43 | 197 | 12 | 9 | 1 | - | - |
| CAMH | 19 | 82 | 7 | 8 | - | 292.35 | ± 302.04 |
| CASSI | 0 | 46 | 3 | 4 | - | 598.24 | ± 502.88 |
| COBRE | 0 | 59 | 6 | 1 | 3 | 433.53 | ± 390.65 |
| EONCKS | - | - | - | - | 90 | 1367.94 | ± 1667.11 |
| ESO | 0 | 27 | 0 | 3 | 4 | 385.07 | ± 312.51 |
| FBIRN | 0 | 136 | 19 | 10 | 18 | 373.95 | ± 387.19 |
| FIDMAG | 2 | 100 | 9 | 27 | 22 | 573.75 | ± 485.16 |
| FSLRome | 10 | 81 | 26 | 42 | 3 | 305.14 | ± 229.17 |
| GROUP | - | - | - | - | 85 | - | - |
| Huilong | 0 | 93 | 7 | 0 | 134 | - | - |
| IGP | 0 | 2 | 37 | 2 | 5 | 654.07 | ± 988.48 |
| IMH | 0 | 66 | 60 | 24 | 1 | 200.23 | ± 183.66 |
| iRELATE | 0 | 0 | 0 | 49 | 1 | 927.4 | ± 2114.38 |
| JBNU | 28 | 62 | 1 | 3 | - | 351.08 | ± 232.14 |
| Madrid | 1 | 10 | 0 | 0 | - | - | - |
| MCIC | 10 | 8 | 0 | 124 | 6 | 533.53 | ± 581.95 |
| MPRC | - | - | - | - | 199 | - | - |
| PAFIP | 0 | 319 | 24 | 0 | - | 195.2 | ± 81.17 |
| RSCZ | 0 | - | - | - | 40 | - | - |
| SCORE | 37 | 31 | 1 | 0 | - | 230.62 | ± 298.57 |
| SNUH | 9 | 22 | 0 | 1 | - | 167.88 | ± 186.11 |
| SWIFT | 0 | 18 | 2 | 0 | 4 | 553.95 | ± 269.94 |
| UCISZ | - | - | - | - | 27 | - | - |
| UNIBA | 0 | 33 | 1 | 6 | 21 | 180.97 | ± 94.52 |
| Zurich | 3 | 55 | 0 | 2 | - | 494.76 | ± 456.67 |
| NA: this information was either not available or not applicable to N number of participants (per cohort) | | | | | | | |

**Supplementary Table S3.** ENIGMA – Schizophrenia Working Group instrument for diagnosing schizophrenia and exclusion criteria by cohort.

| Cohort | Country | Diagnosis measurement | Sample characteristics/inclusion criteria | Exclusion criteria |
|---|---|---|---|---|
| ASRB | Australia | Diagnosis was confirmed using the OPCRIT algorithm applied to interviewer ratings on the DIP, acc. to ICD-10 criteria | All participants were fluent English speakers and aged 18-65 years old | No history of an organic brain disorder, brain injury accompanied by > 24 h of amnesia, mental retardation defined as an IQ < 70, movement disorder, current substance dependence, or electro-convulsive therapy in the preceding 6 months. The control participants additionally had no personal history of psychotic disorder or family history of psychotic disorder in their first-degree biological relatives. |
| CAMH | Canada | SCID DSM-IV-TR Axis I | Schizophrenia outpatients who were clinically stable as determined by no medication change within the past month | Exclusion criteria were intelligence quotient < 70 as estimated by the Wechsler Test for Adult Reading (WTAR), substance dependence or abuse reported or indicated by a urine toxicology screen, head trauma with loss of consciousness, neurological disorders, and any magnetic resonance imaging contraindications. A first degree relative with a primary psychotic disorder was also an exclusion criterion for controls. |
| CASSI | Australia | SCID DSM-IV-TR AXIS I | A diagnosis of schizophrenia or schizoaffective disorder; all patients were between 18 and 51 years of age and had been receiving antipsychotics for at least 1 year before entering the study | Patients with a concurrent Axis I psychiatric diagnosis, a history of substance abuse or dependence (within the past 5 years), head injuries with loss of consciousness, seizures, central nervous system infection, untreated diabetes or hypertension, mental retardation or contraindications to the administration of raloxifene were excluded. Women were excluded if they were currently pregnant or were receiving hormone therapy and refused alternate forms of birth control. For the healthy controls, exclusion criteria consisted of a personal history of or a first-degree relative with a DSM-IV Axis I psychiatric diagnosis, history of substance abuse or dependence (within the past 5 years), head injuries with loss of consciousness, seizures, central nervous system infection, untreated diabetes or hypertension or mental retardation. Any participants having excessive motion or other artifacts and/or clinically relevant neuroanatomical abnormalities were excluded. |

| | | | | |
|---|---|---|---|---|
| COBRE | USA | SCID DSM-IV Axis I Disorders | All participants were in the 18-65 age range and had a diagnosis of schizophrenia. Healthy individuals were included if they did not have a personal or family history of psychiatric disorders. | History of neurological disorder, history of mental retardation, history of severe head trauma with more than 5 minutes loss of consciousness, history of substance abuse or dependence within the last 12 months and MRI contraindications. |
| EONCKS | South Africa | DSM-IV TR | Men and women; in- or out-patients; aged 16 to 45 years; experiencing a first psychotic episode; and meeting diagnostic criteria for schizophrenia, schizophreniform or schizoaffective disorder. A group of healthy controls, matched for age, gender, ethnicity and educational status was recruited from the same catchment area as the patient group. | Exclusion criteria were a lifetime exposure to antipsychotic medication for longer than 4 weeks; any serious or unstable general medical condition; substance-induced psychosis; and an educational level of lower than Grade 7. Controls were excluded if they met the criteria for any past or current psychiatric disorder. |
| ESO | Czech Republic | ICD-10 (F20.x, F23, F25) | Early or first-episode psychosis, Czech language as a mother tongue, 18-60 years old | Neurocognitive disorders (organic mental disorder), mental disorders caused by addiction, mental retardation (IQ<80), severe neurological disorder, head injury, hypertension, cerebrovascular disease, epilepsy, migraine, endocrine disorders. |
| FBIRN | USA | SCID DSM-IV-TR | All subjects diagnosed with schizophrenia were clinically stable outpatients whose antipsychotic medications and doses had not changed within the last two months. | Schizophrenia and healthy volunteers with a history of major medical illness, drug dependence in the last five years (except for nicotine), current substance abuse disorder, or MRI contraindications, were excluded. Individuals with schizophrenia who had significant tardive dyskinesia and healthy volunteers with a current or past history of major neurological or psychiatric illness or with a first-degree relative with a DSM IV Axis-I psychotic disorder diagnosis were also excluded. |
| FIDMAG | Spain | DSM-IV criteria based on interview and review of clinical history | Patients had a diagnosis of schizophrenia. All participants were in the 18-65 age range. | Controls were excluded if they reported a history of mental illness and/or treatment with psychotropic medication. Patients were excluded if have had a history of brain trauma or neurological disease or had shown alcohol/ substance abuse within 12 months before participation |
| FSLRome | Italy | SCID DSM-IV Axis I Disorders (SCID-I) and SCID DSM-IV Axis II Personality Disorders (SCID-II) | Inclusion criteria were (i) age between 18 and 65 years; (ii) at least five years of education; and (iii) suitability for MRI scanning. | Exclusion criteria were (i) history of alcohol or drug abuse in the two years before the assessment; (ii) lifetime drug dependence; (iii) traumatic head injury with loss of consciousness; (iv) past or present major medical illness or neurological disorders; (v) any (for HC) or additional (for |

| | | | | |
|---|---|---|---|---|
| | | | | patients) psychiatric disorder or mental retardation; (vi) dementia or cognitive deterioration according to DSM-IV-TR criteria, and Mini-Mental State Examination (MMSE) score < 25, consistent with normative data in the Italian population; (vii) not able and willing to give written informed consent. |
| GROUP | The Netherlands | DSM-IV-TR | Patients had a diagnosis of schizophrenia or other non-affective psychotic disorder. Participants were of age between 16 and 50. | (iv) not able and willing to give written informed consent. |
| Huilong | China | DSM-IV criteria | Patients who met criteria for schizophrenia. All participants were Han Chinese and were recruited from Beinjing Huilonguan Hospital. | No current or past neurological conditions, unstable major medical conditions, or current or previous substance dependence (although nicotine dependence was permitted) |
| IGP | Australia | Diagnosis was confirmed using the OPCRIT algorithm applied to interviewer ratings on the DIP, acc. to ICD-10 criteria | All participants were fluent English speakers and aged 18-65 years old | General exclusion criteria included an inability to communicate sufficiently in English, a current neurological disorder, a diagnosis of substance abuse or dependence in the past six months; and/or having been treated with electroconvulsive therapy in the previous six months. |
| IMH | Singapore | SCID DSM-IV Axis I Disorders | (1) Diagnosis of SZ (Patients), (2) Age: 21-65, (3) English speaking, (4) Provision of informed written consent | History of significant head injury; significant Neurological diseases (such as epilepsy, cerebrovascular accident) or Medical Illnesses; significant DSM IV alcohol or substance use or dependence; contraindications to MRI (e.g. pacemaker, orbital foreign body, recent surgery/procedure with metallic devices/implants deployed); pregnant women; claustrophobia |
| iRELATE | Ireland | SCID DSM-IV | All participants were between the ages of 18 and 65 years and all but three were Caucasian. Healthy participants were included in the study only when they reported no history of psychiatric or general health problems and did not have first-degree relatives with schizophrenia spectrum disorders. | Participants were excluded from the study for the following reasons: (a) a history of acquired brain injury causing loss of consciousness of more than one minute; (b) substance abuse in the preceding six months; (c) IQ < 70; (d) and a diagnosis of a neurological disorder |
| JBNU | South Korea | DSM-Ⅴ | A diagnosis of schizophrenia: all patients were between 18 – 71 years old | Patients with an (a) alcohol or substance use disorder; (b) intellectual disability (IQ ≤70); (c) current or past neurological disease, serious medical illness, or pregnancy; and (d) claustrophobia were excluded. |
| Madrid | Spain | K-SADS acc. to DSM IV TR criteria | Age between 7-18 years; onset of first psychotic positive symptom within a psychotic episode before age of 18; Referral from adolescent inpatient unit (Hospital General Universitario Gregorio Marañón) or local clinical services | Intellectual disability per DSM-IV-TR criteria (IQ < 70 & impaired functioning); pervasive developmental disorder; past history of head trauma with loss of consciousness; pregnancy |

| | | | |
|---|---|---|---|
| | | | (PIENSA program); diagnosis of a psychotic disorder; written informed consent; duration of psychosis at baseline of less than 2 years. |

| | | | | |
|---|---|---|---|---|
| MCIC | USA | SCID DSM-IV (SCID-NP for controls) or CASH were used to diagnose primary and co-morbid psychiatric disorders in controls and patients | All subjects were between the ages of 18 and 60 and spoke English as their native language. To be included in the schizophrenia cohort, patients had to meet diagnostic criteria for schizophrenia, schizoaffective disorder, or schizophreniform disorder. Concerted effort was made to recruit patients early in the course of their illness and especially those who were antipsychotic drug naïve. The healthy control subjects with no current or past history of psychiatric illness including substance abuse or dependence were matched within site to the patient cohort for age, sex, and parental education. Control subjects who had not been diagnosed with any psychiatric disorders, but had been medicated with antidepressants, anti-anxiety medication or medication for sleep disturbance were included in the study provided that the duration of their medication did not exceed 2 months of lifetime use and no medication was used within the 6 months preceding the baseline MRI scan. | Control subjects who met criteria for current or past history of substance abuse or dependence were excluded from the study. Patients, however, were not excluded from the study unless criteria were met for current (i.e., within the past month) abuse or dependence (except for 6 patients who were found to meet criteria for current abuse after the study data was collected). Both patients and controls were excluded if they had (1) an IQ less than 70 based on a standardized IQ test, (2) history of a head injury resulting in prolonged loss of consciousness, neurosurgical procedure, neurological disease, history of skull fracture, severe or disabling medical conditions, or (3) a contraindication for MRI scanning such as pregnancy, metal in body or head including implanted pacemaker, medication pump, vagal stimulator, deep brain stimulator, implanted TENS unit, or ventriculo-peritoneal shunt |

| | | | | |
|---|---|---|---|---|
| MPRC | USA | SCID DSM-IV combined with a review of medical records | Individuals diagnosed with schizophrenia or schizoaffective disorder; and healthy controls without current DSM-IV Axis I psychiatric illnesses | The exclusion criteria included diagnosis with hypertension, hyperlipidemia, type 2 diabetes, heart disorders, major neurologic event such as stroke or transient ischemic attack, and recent substance use disorder (except tobacco and marijuana use). |

| | | | | |
|---|---|---|---|---|
| PAFIP | Spain | SCID DSM-IV for patients confirmed by an independent psychiatrist 6 months after the initial contact. CASH for controls. | Patients had to meet the following criteria: (1) age 15–60 years; (2) living in the catchment area; (3) experiencing a first episode of psychosis; (4) no prior treatment with antipsychotic medication or, if previously treated, a total lifetime of adequate antipsychotic treatment of less than 6 weeks; and (5) meeting DSM-IV criteria for schizophrenia, schizophreniform disorder, brief psychotic disorder, or schizoaffective disorder. | Patients were excluded when meeting DSM-IV criteria for (1) drug dependence (except nicotine dependence), (2) mental retardation, and when having a history of neurological disease or head injury. Controls exclusion criteria were current or past history of psychiatric, neurological or general medical illnesses, including substance dependence and significant loss of consciousness. HCs were selected to have a similar distribution in age, sex, laterality index, drug history and years of education as the patient population. The absence of psychosis in first-degree relatives was also confirmed by clinical records and family interview. After a detailed description of the study, each subject gave written informed consent to participate. |

| | | | | |
|---|---|---|---|---|
| RSCZ | Russian Federation | ICD-10 (F20.x) | Early or first-episode psychosis in-patients (no later than 5 years since the first episode) who were clinically stable and received antipsychotic medication therapy. Mentally healthy controls were recruited from acquaintances of the researchers and clinical staff. All participants were fluent Russian speakers, right-handed males. | Common exclusion criteria for patients and controls were: organic brain disorders, neurological or severe somatic disorders, mental retardation, alcohol or substance abuse, history of head trauma with loss of consciousness for more than 5 min. In addition, controls were excluded if they had a family history of psychiatric illness. |
| SCORE | Switzerland | ICD-10 or DSM-IV criteria | First-episode psychosis patients who fulfilled criteria for brief psychotic disorder. All patients were between 18 and 42 years of age. | History of previous psychotic disorder, psychotic symptoms secondary to an organic disorder, substance abuse (except nicotine), psychotic symptoms associated with an affective psychosis or a borderline personality disorder, age younger than 18 years, inadequate knowledge of the German language, and IQ less than 70 as measured by the Mehrfachwahl Wortschatz Test Form B. |
| SNUH | South Korea | SCID DSM-IV Axis I disorder (SCID-I/SCID-NP for controls) | A diagnosis of schizophrenia, schizophreniform disorder, or schizoaffective disorder; all patients were between 17 and 49 years of age | Common exclusion criteria included substance abuse or dependence (except nicotine), neurological disease or significant head trauma, medical illness that could accompany psychiatric symptoms, and intellectual disability (intelligence quotient [IQ] < 70). |
| SWIFT | Switzerland | SCID DSM-IV-TR | Age between 18-65 y/o, good German language skills that allowed them to understand the consent procedure and to undergo the clinical assessment, right-handedness according to the Edinburgh Inventory. For patients: Diagnosis with a schizophrenia spectrum disorder. | Participants were excluded if they were left-handed, pregnant, showed any contraindications for MRI (e.g. metal-containing implants such as pacemaker or cochlear implants, claustrophobia), had a history of serious neurological issues, or reported current abuse of alcohol and/or psychoactive substances (apart from nicotine). Additionally, controls had no current major psychiatric DSM-IV Axis I diagnoses, as assessed with the screening questionnaire of the Structured Clinical Interview for DSM-IV Axis I Disorders. |
| UCISZ | USA | SCID DSM-IV-TR criteria | All subjects diagnosed with schizophrenia were clinically stable outpatients whose antipsychotic medications and doses had not changed within the last two months. | Schizophrenia and healthy volunteers with a history of major medical illness, drug dependence in the last five years (except for nicotine), current substance abuse disorder, or MRI contraindications, were excluded. Individuals with schizophrenia who had significant tardive dyskinesia and healthy volunteers with a current or past history of major neurological or psychiatric illness or with a first-degree relative with an Axis-I psychotic disorder diagnosis were also excluded.? |
| UNIBA | Italy | SCID DSM-IV | Patients with schizophrenia on stable antipsychotic treatment for at least one month.<br>All subjects age between 18 and 65 years old. | Healthy controls or schizophrenia patients with a history of drug or alcohol abuse within the last 6 months, metal implants or other MRI contraindications, head trauma with loss of consciousness, or any clinically significant medical condition. |

| | | | | |
|---|---|---|---|---|
| Zurich | Switzerland | MINI DSM IV | A diagnosis of schizophrenia | We excluded patients with any other DSM-IV Axis I disorder (in particular, current substance use disorder and major depressive disorder), those medicated with lorazepam at a dose higher than 1 mg, those with florid psychotic symptoms (i.e., any positive subscale item scores higher than 4 on the Positive and Negative Syndrome Scale [PANSS]) and those with extrapyramidal side effects (i.e., a total score higher than 2 on the Modified Simpson–Angus Scale [MSAS]). Healthy controls were screened for any neuropsychiatric disorders using the structured Mini-International Neuropsychiatric Interview to ensure that they had no previous or present psychiatric illness. Both patients and healthy controls were required to have a normal physical and neurologic status and no history of major head injury or neurologic disorder. |

SCID: Structured Clinical Interview for DSM Disorders; CAMH: Comprehensive Assessment of Symptoms and History; MINI DSM IV: Mini-International Neuropsychiatric Interview for DSM IV; OPCRIT: Operational Criteria Checklist for Psychotic Illness and Affective Illness; DIP: Diagnostic Interview for Psychosis; ICD: international classification of disease

**Supplementary Table S4.** Sample image acquisition and image pre-processing details by cohort

| Cohort | Number of scanners | Scanner Vendor & Type | Imaging Protocols | Slice orientation | FreeSurfer Version | Operating System | Number of subjects removed from analysis due to QC failure |
|---|---|---|---|---|---|---|---|
| ASRB | 5 | Siemens Avanto 1.5T | High-resolution T1-weighted structural magnetic resonance imaging (sMRI) brain scans (MPRAGE) were acquired using an optimized magnetization prepared rapid acquisition gradient echo on 1.5 T Siemens Avanto scanners (Siemens, Erlangen, Germany) across five Australian research sites (Loughland and al., 2010). Image parameters were set to 176 slices of 1mm thickness, no gap with field-of-view 250 x 250 mm2, repetition time 1980 ms, echo time 4.3 ms, data acquisition matrix 256 x 256, with a flip matrix of 15°, resulting in a voxel size of 0.98×0.98×1.0 mm3 | Sagittal | v5.1.0 | Mac OSX | 0 |
| CAMH | 1 | GE 1.5T | SPGR, TR/TE/TI=12.3/5.3/300ms, flip angle=20°, 256x256x128 matrix, FOV=240x240mm, slice thickness=1.5mm | Axial | v5.3.0 | xubuntu x86_64-linux | 0 |
| CASSI | 1 | 3-Tesla Philips Achieva with an 8-channel head coil | T1 weighted high-resolution anatomical scans were obtained (1mm slice thickness, no gap, 180 slices, TR = 5.4ms, TE = 2.4ms, field of view 256mm) | Sagittal | v.5.1.0 | Mac OSX 10.8 | Scans from three participants were excluded for motion artifact (1 control and 2 patients), and two participants due to neuroanatomical abnormalities (one healthy control showing a meningioma and one patient with agenesis of the corpus callosum) |
| COBRE | 1 | 3T Siemens TIM Trio | T1-weighted images were acquired with a 5-echo multi-echo MPRAGE sequence [TE (echo times) = 1.64, 3.5, 5.36, 7.22, 9.08 ms, TR (repetition time) = 2.53 s, TI (inversion time) = 1.2 s, 7○ flip angle, number of excitations (NEX) = 1, slice thickness = 1 mm, FOV (field of view) = 256 mm, resolution = 256x256] | Sagittal | v5.3.0 | Linux RedHat | A total of 9 participants were excluded following ENIGMA QA protocol. |
| EONCKS | 1 | 3T siemens allegra | MPRAGE sequence, 2080 ms repetition time; 4.88 ms echo-time, Field of view: 230 mm, 176 slices, 0.9 × 0.9 × 1 mm3 voxel size | Sagittal | v 6.0 | Linux | Nineteen patients and 11 controls were excluded due to motion artefacts or a technical scanner error. |

| | | | | | | | |
|---|---|---|---|---|---|---|---|
| ESO | 1 | 3T Siemens Tim Trio | MP-RAGE 3D, 1mm thickness, acquisition matrix 256 x 256, TR=2300ms, TE=4.63ms,TI=900ms | Sagittal | v5.3.0 | Linux | Only subjects without significant motion artifacts (assessed by visual inspection) were included. Apart from ENIGMA QA protocol, visual inspection of all slices and edits to the skullstrip, white matter segmentation and control points insertion for correction of signal intensity normalization were done where needed. |
| FBIRN | 7 | 3T Siemens Tim Trio; 3T GE | High-resolution structural imaging scans were acquired on six 3T Siemens Tim® Trio System and one 3T General Electric Discovery MR750 scanner. MP-RAGE scan parameters for the Siemens scanner were: scan plane=sagittal, TR/TE/TI=2300/2.94/1100ms, GRAPPA acceleration factor=2, flip angle=9°, resolution=256×256x160, FOV=220mm2, voxel size=0.86x0.86x1.2mm, and NEX=1. IR-SPGR scan parameters for the General Electric scanner were: scan plane=sagittal, TR/TE/TI=5.95/1.99/450ms, ASSET acceleration factor=2, a flip angle=12°, resolution=256×256x166, FOV=220mm2, voxel size=0.86x0.86x1.2mm, and NEX=1. All scans covered the entire brain. | Sagittal | v5.3.0 | Centos 64bit | 0 |
| FIDMAG | 1 | 1.5T GE Signa | 180 axial slices; 1mm slice thickness, no gap, matrix size 512x512; 0.5x0.5x1mm3 voxel resolution; TE 4ms, TR 2000ms, flip angle 15 | Axial | v5.3.0 | Linux Ubuntu | 0 |
| FSLRome | 1 | Siemens 3T Allegra | 3D MPRAGE: TE/TR=2.4/7.92 ms, flip angle=15°, voxel size 1×1×1 mm | Sagittal | 6.0dev | Linux | |
| GROUP | 1 | Siemens 3T Allegra | MDEFT; 176 slices, 1 mm isotropic voxel size, TR/TE/TI=7.92/2.4/910ms, flip angle 15°, total acquisition time 12 min and 51 s<br>MPRAGE; 192 slices, 1 mm isotropic voxel size, TR/TE/TI 2250/2.6/900ms, flip angle 9°, total acquisition time 7 min and 23 s. The matrix size was 256 × 256 and field of view was 256 × 256 mm2. The number of excitations was one. | Sagittal | v5.0.0 | Mac OSX | 0 |
| Huilong | 2 | Huilong 1: 3T Siemens | Huilong 1: T1-weighted, 3D MPRAGE, 1x1x1mm, TE/TR/TI=.9/2300/900ms, flip angle=9 degrees. | Sagittal | v5.3.0 | Linux | |

| | | | | | | | |
|---|---|---|---|---|---|---|---|
| | | Verio<br><br>Huilong 2: 3T GE Signa HDxt | Huilong 2: T1-weighted, 3D BRAVO, 1x1x1mm, TE/TR/TI=2.5/6.8/1100ms, flip angle=7 degrees. | | | | |
| IGP | 1 | Philips 3T Achieva TX | TR 8.9ms, TE 4.1ms, field of view 240mm, matrix 268 x 268, 200 sagittal slices, slice thickness 0.9mm, no gap | Sagittal | v5.3.0 | Mac OSX | 0 |
| IMH | 1 | 3T Philips Achieva | T1 scans: 180 axial slices of 0.9mm thickness with no gap, FOV = 230x230 mm2, matrix 256x204, voxel size =0.89x0.89x0.9 mm3, TR=7.2 s, TE=3.3 ms, FA=8°, | Axial | v5.3.0 | Mac OSX | 2 due to excessive motion artifacts |
| iRELATE | 1 | 3T Philips Achieva | T1 scans: FFE pulse sequence, TR/TE = 8.5/3.9 ms, FOV = 256 × 256 × 160 mm, voxel size=1x1x1mm, TI (Inversion Time) = 1060 ms, flip angle = 8°, SENSE factor = 1.5 and acquisition time = 7 min 30 s) | Sagittal | V6.0.0 | Linux | Total 5, Sz=3, HC=2 |
| JBNU | 1 | 3T<br><br>Siemens MAGNETOM Verio syngo | T1-weighted, 3-dimensional Magnetization Prepared Rapid Gradient Echo (TR = 1900ms; TE=2.45ms; FOV=250mm; FA=9°; voxel size=1x1x1mm$^3$; Slice thickness=1mm ) | Sagittal | V6.0.0 | Ubuntu<br><br>18,04.5 LTS (x86_64) | 0 |
| Madrid | 1 | Philips Intera 1.5T | T1 scans; TR 25ms; TE 9.18ms; voxel size 1x0.94x0.94mm$^3$; FOV 240x240m$^2$; 175 slices; flip angle 30; Gradient Echo (FFE 3d) | Sagittal | v5.3.0 | Linux | 0 |
| MCIC | 3 | 1.5, 3T Siemens and GE | T1 scans: TR = 2530 ms for 3 T, TR = 12 ms for 1.5 T; TE = 3.79 ms for 3 T, TE = 4.76 ms for 1.5 T; FA = 7 for 3 T, FA = 20 for 1.5 T; TI = 1100 for 3 T; Bandwidth = 181 for 3 T, Bandwidth = 110 for 1.5 T; 0.625×0.625 mm voxel size; slice thickness 1.5 mm; FOV 256×256×128 cm matrix; FOV = 16 cm (could be increased to 18 cm when needed for full brain coverage). | Coronal | v4.0.1 | Linux of various flavors | 5 subjects failed automated segmentation procedure due to excessive motion artifacts 2 participants' MRI data failed the manual inspection |
| MPRC | 2 | MPRC 1: 3T Siemens Allegro | MPRC 1: T1-weighted, 3D MPRAGE, 1x1x1mm, TE/TR/TI=4.3/2500/1000ms, flip angle=8 degrees.<br><br>MPRC 2**:** T1-weighted, 3D MPRAGE, 1x1x1mm, | Sagittal | v5.3.0 | | |

| | | | | | | | |
|---|---|---|---|---|---|---|---|
| | | MPRC 2: 3T Siemens Trio | TE/TR/TI=2.9/2300/900ms, flip angle=9 degrees. | | | Linux | |
| PAFIP | 2 | GE 1.5T | Three-dimensional T1-weighted images, using a spoiled grass (SPGR) sequence acquired in the coronal plane with: echo time (TE)=5 ms, repetition time (TR)=24 ms, numbers of excitations (NEX)=2, rotation angle=45°, field of view (FOV)=26×19.5 cm, slice thickness=1.5mm and a matrix of 256×192. | Coronal | v5.0.0 | Ubuntu 11,04 (x86_64) | 1 subject was excluded because motion artifacts resulted in very poor segmentation |
| RSCZ | 1 | 3T Philips Achieva | A turbo field echo sequence covering the whole brain. TR = 8.2 ms, TE = 3.7 ms, flip angle = 8, FOV = 240 mm, voxel size of 0.83 × 0.83 mm with a slice thickness of 1 mm, no gap. | Sagittal | v5.3.0 | Centos 6.6 | 0. Only subjects without significant motion and other artifacts (assessed by visual inspection) were included for carrying out ENIGMA QA protocol. |
| SCORE | 1 | 3T Philips Achieva | MPRAGE: aquicition matrix: 256×256×176, isotropic spatial resolution: 1x1x1mm3, TI=1000ms, TR=2s, TE=3.4 ms, flip angle: 8° and bandwidth of 200 Hz/pixel | Sagittal | 6.0dev | ubuntu 18.04 LTS | From the initial data set, 11 subjects had to be excluded based on erroneous brain segmentation. Among them, 4 subjects revealed statistical outliers. |
| SNUH | 1 | 3T Siemens Trio | high-resolution T1-weighted, threedimensional Magnetization Prepared Rapid Gradient Echo (TR = 670ms; TE=1.89ms; FOV=250mm; FA=9°; voxel size=1x1x1mm3) | Sagittal | v5.3.0 | OSX 10.9 | 0 |
| SWIFT | 1 | 3T Siemens Verio | MPRAGE: 160 sagittal slices, 1mm slice thickness, 256x256 matrix size, 1x1x1 mm3 voxel size. TR = 2.3ms, TE = 2.98ms, TI = 900ms. | Sagittal | v 7.1.1 | Linux | 0 |
| UCISZ | 1 | 3T Philips Achieva | T1TFE:200 sagittal slices, 320x274 matrix size, .75mm isotropic, TR = 11ms, TE=4.562ms, flip angle = 18°, | Sagittal | v6.0dev | Centos 3.10.72-1.el6.elrepo.x86_64 | 0 |
| UNIBA | 1 | 3T GE | T1-weighted 3D FFE, TR/TE 9.86/4.6ms, 0.875x0.875x1 voxels, flip angle 8, FOV 224x160x168, 160 slices | Axial | v5.3.0 | Ubuntu 10.04, Kernel Linux 2.6.32-25-generic, GNOME 2.30.2 | 3 |
| Zurich | 1 | 3T Philips | 3D T1-weighted images were acquired with an ultra fast gradient echo T1-weighted sequence(TR=8.4ms, TE=3.8ms, flip angle=8°) in 160 sagittal plan slices | Sagittal | v6.0.0 | Linux | 0 |

| | | | |
|---|---|---|---|
| | | | (1mm slice thickness, no slice gap) of 240×240mm2 resulting in 1x1x1mm3voxels. |

**Supplementary Table S5.** Additional results for antipsychotic use and brain-PAD in SZ

| Clinical parameter | N | K | beta | SE | 95% CI | p-value |
|---|---|---|---|---|---|---|
| AP user – typical vs. atypical | 789 (116/673) | 5 | 0.18 | 0.83 | -1.48, 1.84 | 0.83 |
| AP user – both vs. atypical | 275 (235/40) | 3 | 0.53 | 1.15 | -1.72, 2.78 | 0.64 |

Additional results for the effects of antipsychotic use on brain-PAD scores (predicted brain age – chronological age) in SZ. The regression coefficient *beta* indicates a mean brain-PAD difference between typical, or both typical and atypical, versus atypical antipsychotic use. Effects were adjusted for age and age$^2$. N: total number of participants included in each analysis (total group size for type of AP use is given in brackets); K: number of cohorts; SE: standard error; AP: Antipsychotics; CI: confidence interval.

**Supplementary Table S6.** Clinical characteristics and brain-PAD in SZ, additionally adjusting for handedness

| Clinical parameter | N | K | beta | SE | 95% CI | p-value |
|---|---|---|---|---|---|---|
| Age of onset (years) | 1783 | 16 | -0.02 | 0.09 | -0.20, 0.17 | 0.85 |
| Length of illness (years) | 1785 | 16 | 0.01 | 0.09 | -0.17, 0.19 | 0.94 |
| PANSS total | 1099 | 14 | 0.05 | 0.08 | -0.08, 0.18 | 0.44 |
| SANS global | 1539 | 15 | 0.17 | 0.12 | -0.05, 0.40 | 0.13 |
| SAPS global | 1152 | 15 | 0.26 | 0.14 | -0.11, 0.42 | 0.21 |
| AP user – atypical vs. unmed | 572 (453/119) | 6 | 2.22 | 1.49 | -0.70, 5.15 | 0.14 |
| AP user – typical vs. unmed | 114 (42/72) | 3 | 0.07 | 1.01 | -1.91, 2.05 | 0.95 |
| AP user – both vs. unmed | 261(179/82) | 4 | -0.11 | 1.05 | -2.17, 1.95 | 0.92 |
| AP user – typical vs. atypical | 675 (108/567) | 4 | 0.18 | 1.07 | -1.91, 2.28 | 0.86 |
| AP user – both vs. atypical | 275 (40/235) | 3 | 0.53 | 1.15 | -1.72, 2.78 | 0.64 |
| CPZ-equivalent dose | 1424 | 14 | 0.00 | 0.01 | -0.02, 0.03 | 0.83 |

Association of clinical characteristics and brain-PAD (predicted brain age – chronological age) in SZ, after additional adjustment for handedness. For continuous variables (age of onset, length of illness, PANSS total and CPZ), the regression coefficient *beta* indicates a mean change in brain-PAD per unit increase in each clinical variable across cohorts. For categorical variables (AP use – typical/atypical or both atypical and typical), beta indicates the mean brain-PAD difference between each treatment group and unmedicated individuals or atypical AP use. Effects were adjusted for handedness, age and age$^2$. P-values were not corrected for multiple comparisons. K: number of cohorts; N: total number of participants included in each meta-analysis (where applicable, total group size for AP type use/unmedicated or AP type use/atypical AP use is given in brackets); SE: standard error; CI: confidence interval; SANS: Scale for the Assessment of Negative Symptoms; SAPS: Scale for the Assessment of Positive Symptoms; PANSS: Positive and Negative Syndrome Scale; AP: Antipsychotics; CPZ: chlorpromazine.

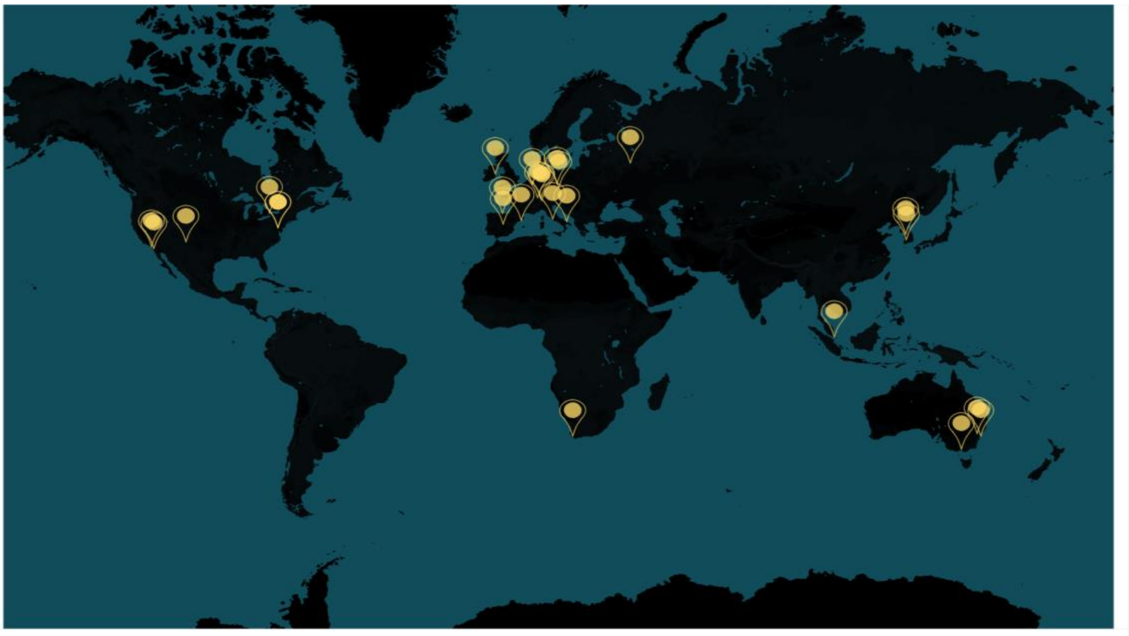

**Supplementary Figure S1.** ENIGMA Schizophrenia Working group world map showing the location of each sample included in this study.

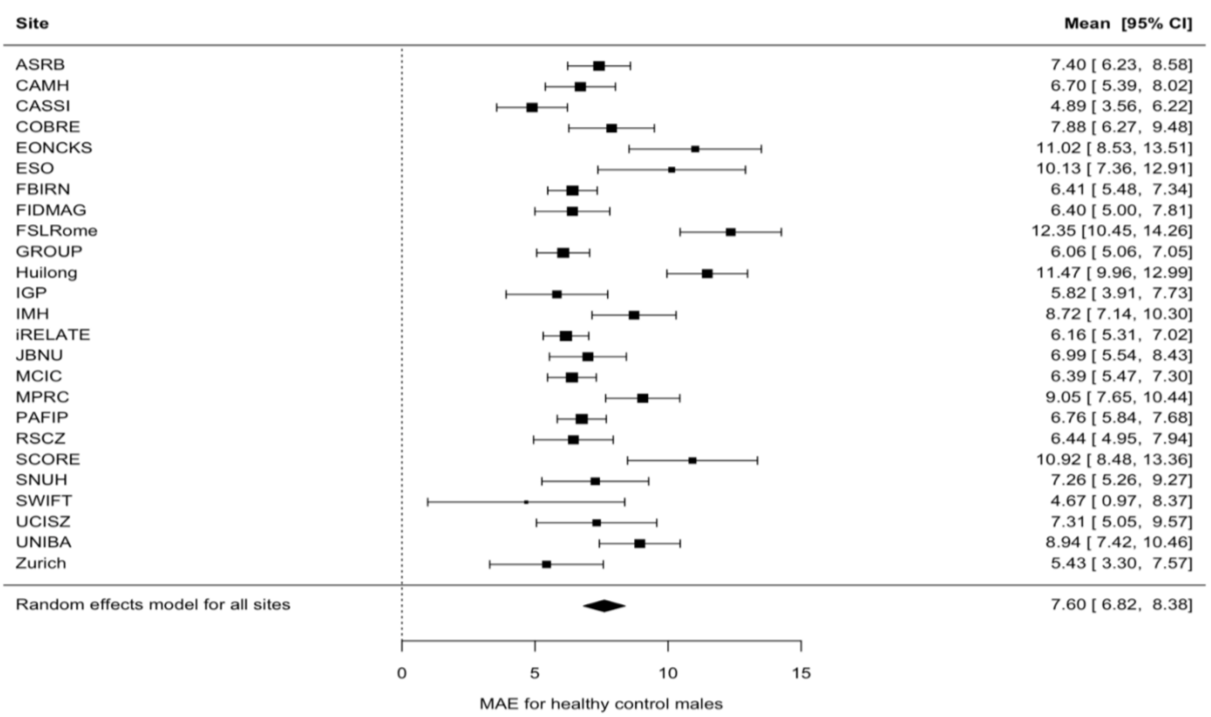

**Supplementary Figure S2a.** Mean absolute error for healthy control males**.** Forrest plot of mean absolute error (MAE) for healthy control males across 25 cohorts. MAE for each cohort is denoted by black boxes and black lines indicate 95% confidence intervals. The size of the box indicates the weight the cohort received (based on inverse variance weighting). The pooled estimate for all cohorts is represented by a black diamond, with the outer edges of the diamond indicating the confidence interval limits.

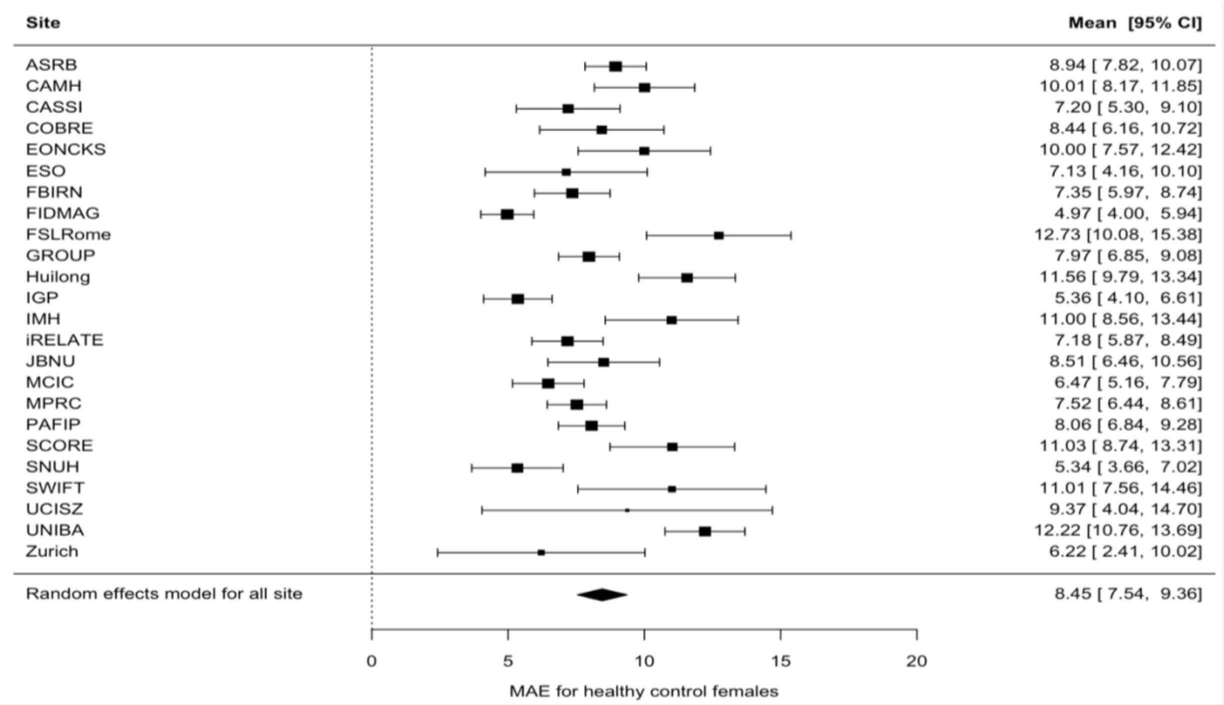

**Supplementary Figure S2b.** Mean absolute error for healthy control females**.** Forrest plot of mean absolute error (MAE) for healthy control females across 24 cohorts. MAE for each cohort is denoted by black boxes and black lines indicate 95% confidence intervals. The size of the box indicates the weight the cohort received (based on inverse variance weighting). The pooled estimate for all cohorts is represented by a black diamond, with the outer edges of the diamond indicating the confidence interval limits

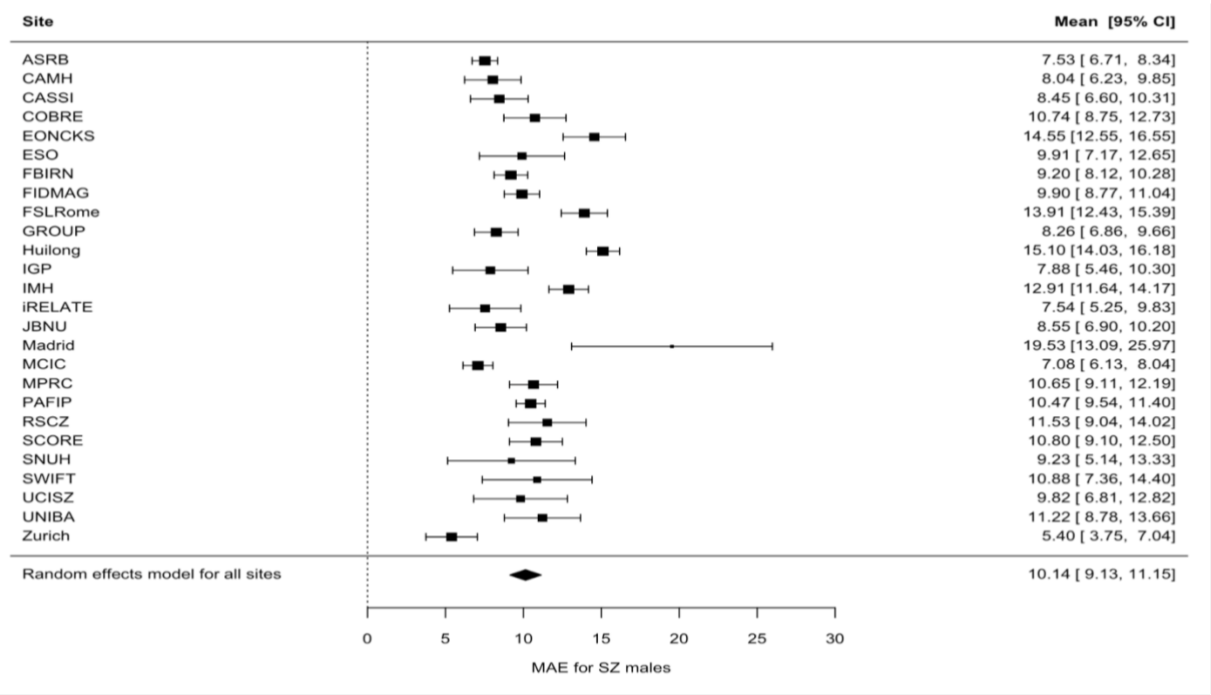

**Supplementary Figure S2c.** Mean absolute error for schizophrenia males. Forrest plot of mean absolute error (MAE) for schizophrenia (SZ) males across 26 cohorts. MAE for each cohort is denoted by black boxes and black lines indicate 95% confidence intervals. The size of the box indicates the weight the cohort received (based on inverse variance weighting). The pooled estimate for all cohorts is represented by a black diamond, with the outer edges of the diamond indicating the confidence interval limits

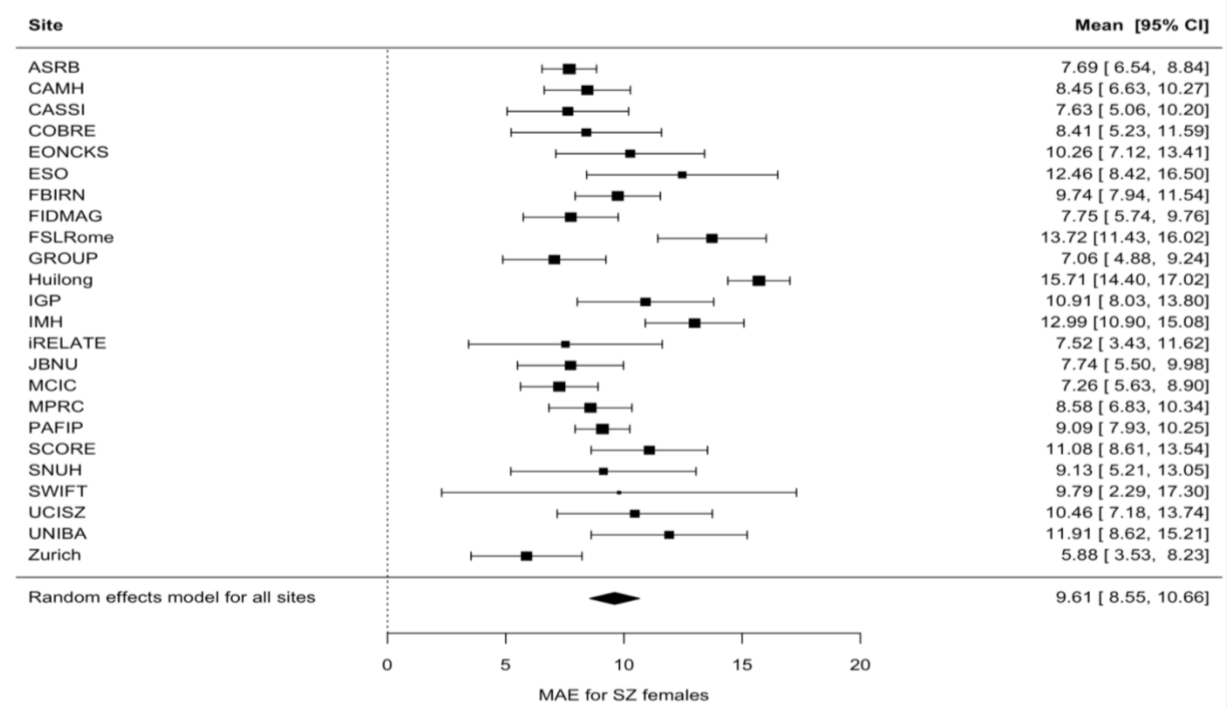

**Supplementary Figure S2d.** Mean absolute error for schizophrenia females. Forrest plot of mean absolute error (MAE) for schizophrenia (SZ) females across 24 cohorts. MAE for each cohort is denoted by black boxes and black lines indicate 95% confidence intervals. The size of the box indicates the weight the cohort received (based on inverse variance weighting). The pooled estimate for all cohorts is represented by a black diamond, with the outer edges of the diamond indicating the confidence interval limits

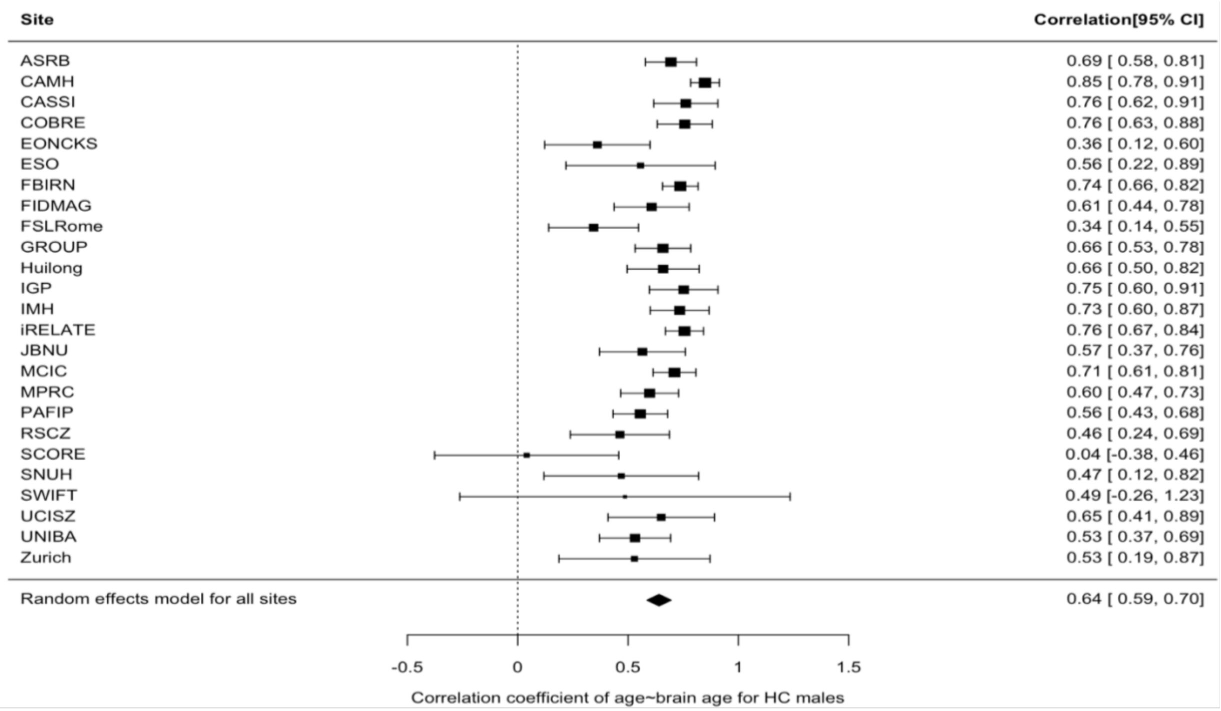

**Supplementary Figure S3a.** Correlation between brain-predicted age and chronological age in healthy control males**.** Forrest plot of correlation between brain-predicted age and chronological age in healthy control males across 25 cohorts. Pearson's correlation coefficient for each cohort is denoted by black boxes and black lines indicate 95% confidence intervals. The size of the box indicates the weight the cohort received (based on inverse variance weighting). The pooled estimate for all cohorts is represented by a black diamond, with the outer edges of the diamond indicating the confidence interval limits.

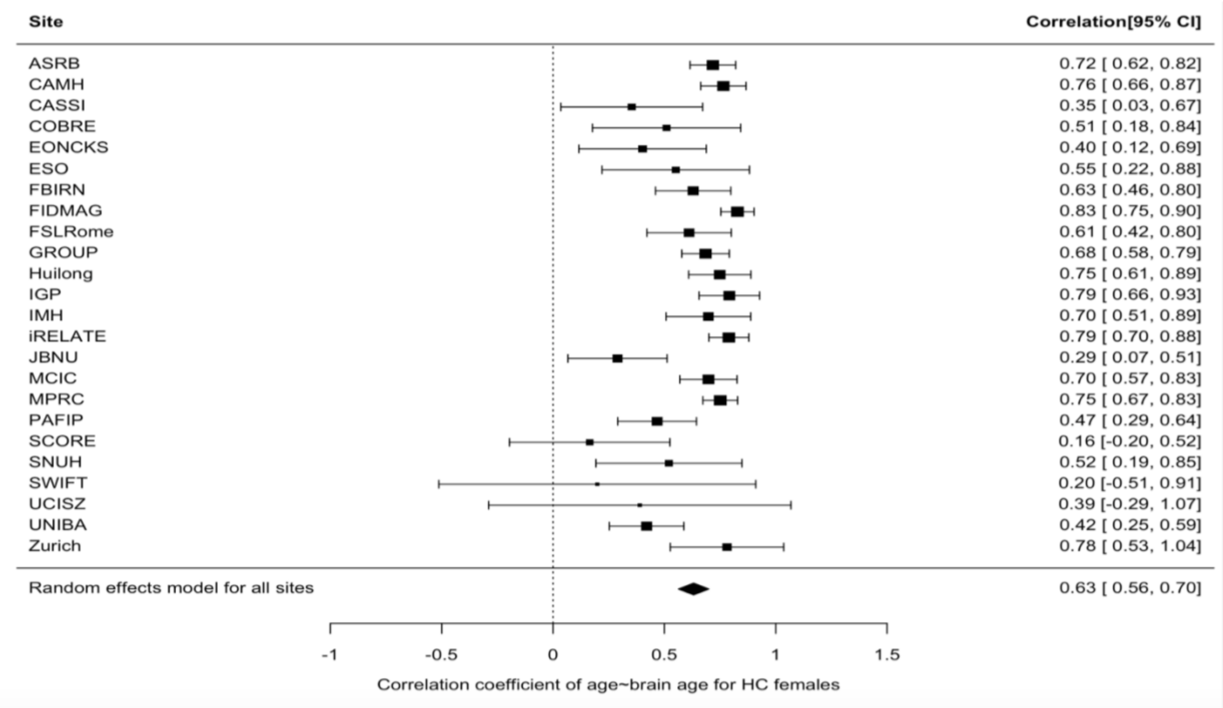

**Supplementary Figure S3b.** Correlation between brain-predicted age and chronological age in healthy control females. Forrest plot of correlation between brain-predicted age and chronological age in healthy controls females across 24 cohorts. Pearson's correlation coefficient for each cohort is denoted by black boxes and black lines indicate 95% confidence intervals. The size of the box indicates the weight the cohort received (based on inverse variance weighting). The pooled estimate for all cohorts is represented by a black diamond, with the outer edges of the diamond indicating the confidence interval limits.

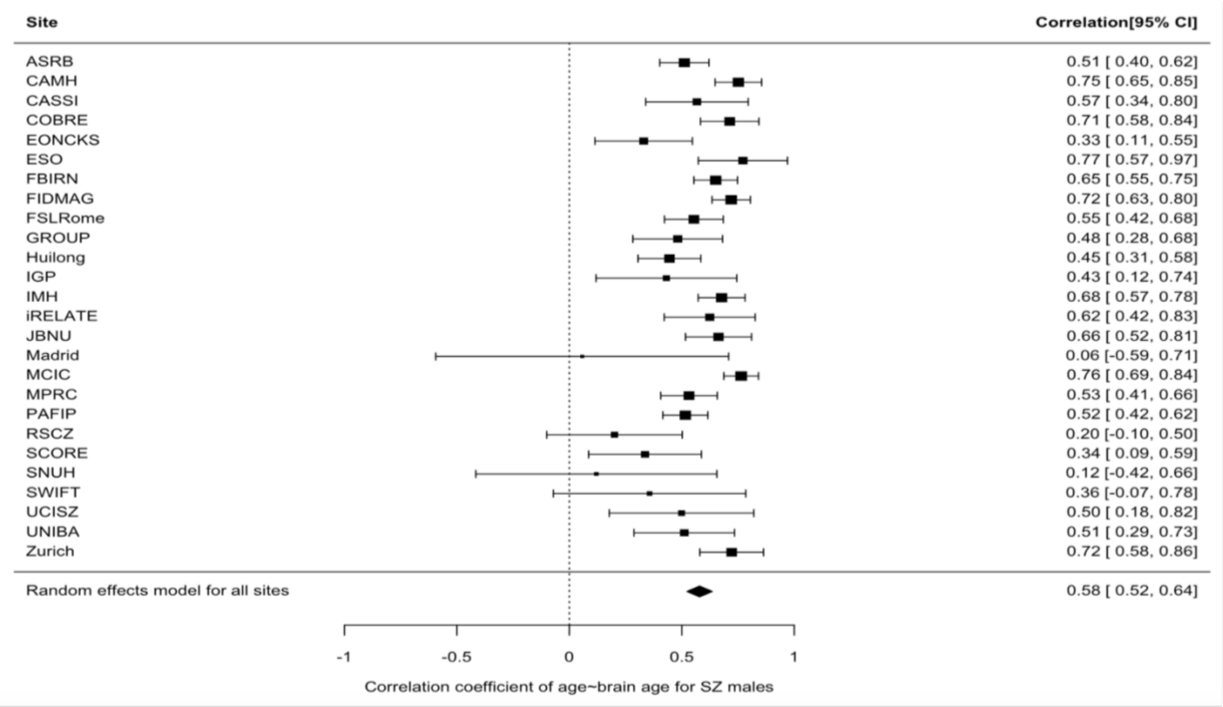

**Supplementary Figure S3c.** Correlation between brain-predicted age and chronological age in schizophrenia males. Forrest plot of correlation between brain-predicted age and chronological age in schizophrenia (SZ) males across 26 cohorts. Pearson's correlation coefficient for each cohort is denoted by black boxes and black lines indicate 95% confidence intervals. The size of the box indicates the weight the cohort received (based on inverse variance weighting). The pooled estimate for all cohorts is represented by a black diamond, with the outer edges of the diamond indicating the confidence interval limits.

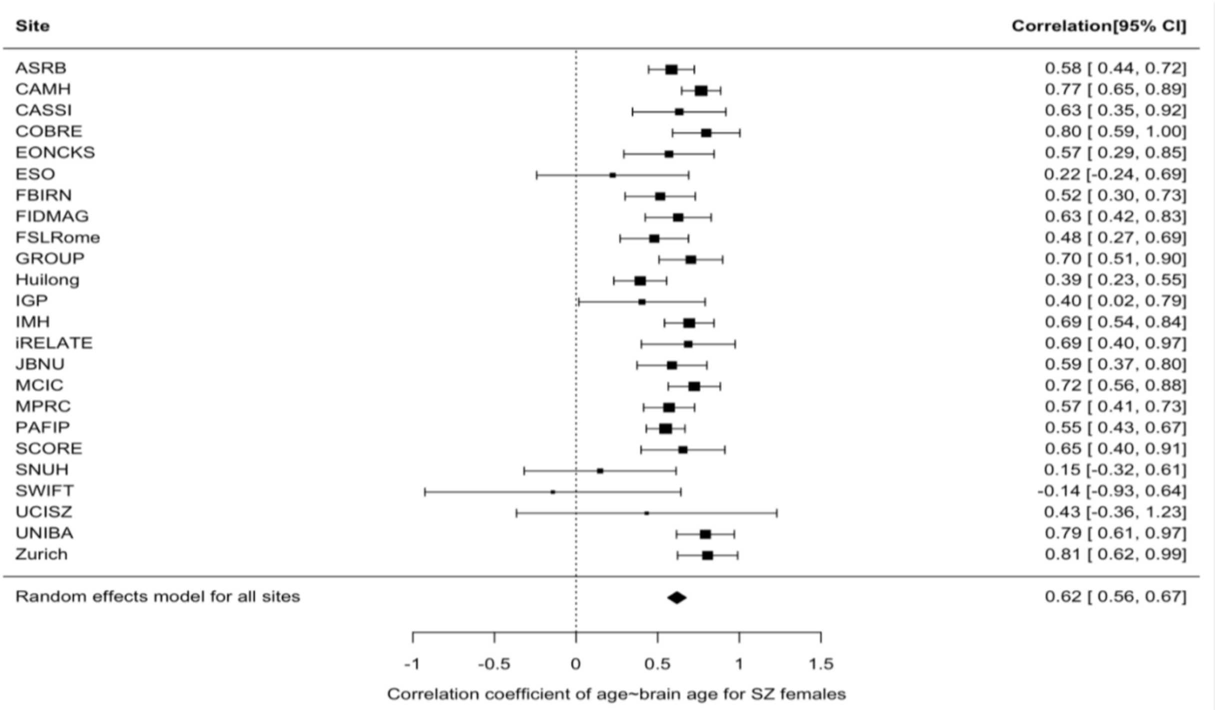

**Supplementary Figure S3d.** Correlation between brain-predicted age and chronological age in schizophrenia females. Forrest plot of correlation between brain-predicted age and chronological age in schizophrenia (SZ) females across 24 cohorts. Pearson's correlation coefficient for each cohort is denoted by black boxes and black lines indicate 95% confidence intervals. The size of the box indicates the weight the cohort received (based on inverse variance weighting). The pooled estimate for all cohorts is represented by a black diamond, with the outer edges of the diamond indicating the confidence interval limits.

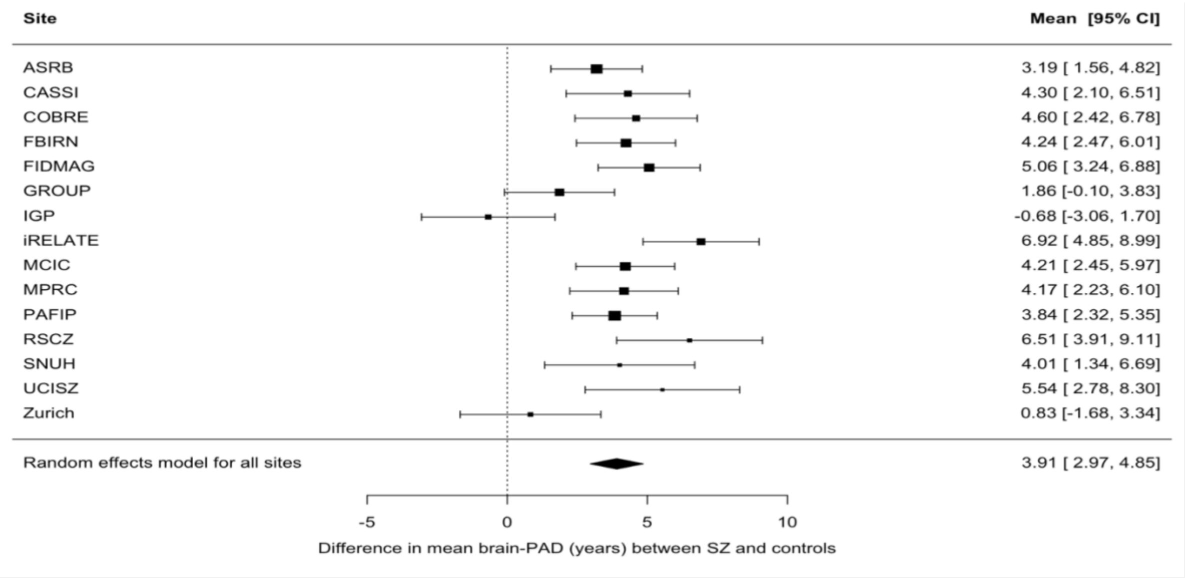

**Supplementary Figure S4.** Sensitivity analysis of case-control differences in brain-PAD. Forest plot of differences in mean brain-PAD scores (predicted brain age - chronological age) between patients with schizophrenia (SZ) and controls across 15 cohorts ($N_{SZ}$ = 1757; $N_{HC}$ = 1712), after excluding 10 cohorts for which the model generalised relatively less well in either male or female health controls - here defined as an MAE > 10.00 or $R^2$ < 0.1. Regression coefficients (in years) are denoted by black boxes. Black lines indicate 95% confidence intervals. Effect sizes were adjusted for sex, age and age$^2$. The size of the box indicates the weight the cohort received (based on inverse variance weighting). The pooled estimate for all cohorts is represented by a black diamond, with the outer edges of the diamond indicating the confidence interval limits.

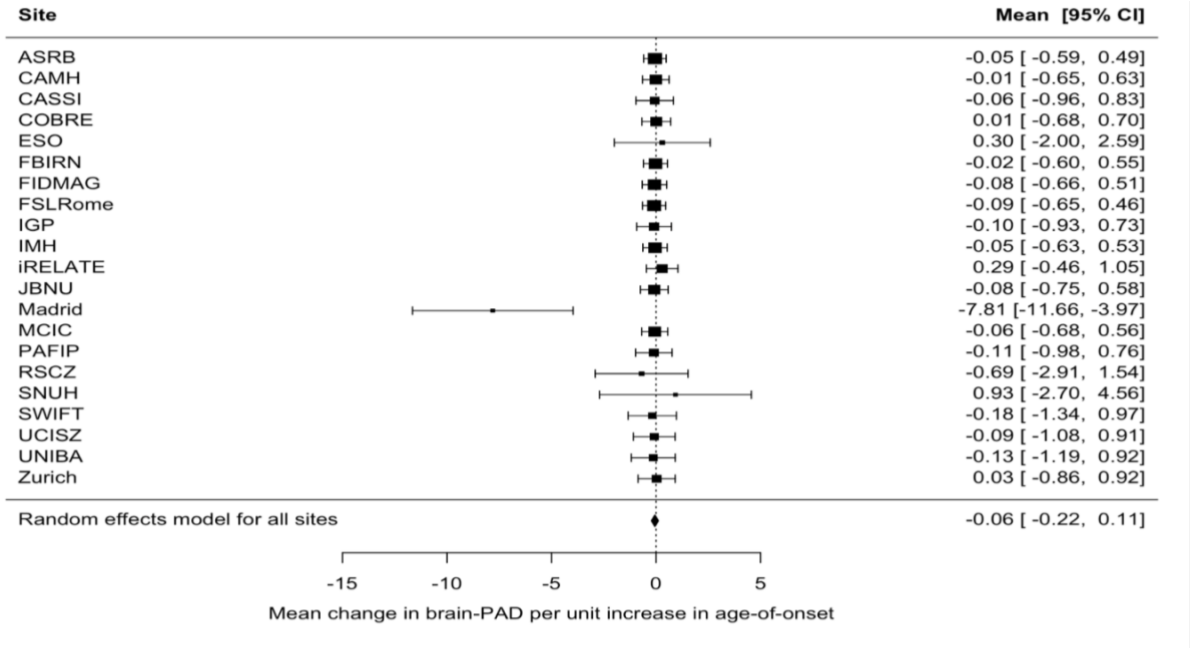

**Supplementary Figure S5a.** Age of illness onset and brain-PAD. Forest plot of the association between age of onset and brain-PAD in SZ across 21 cohorts (N=2053). Regression coefficients are denoted by black boxes and indicate a mean change in brain-PAD (years) per unit increase in age of onset (years), after adjusting for age and age$^2$. Black lines indicate 95% confidence intervals. The size of the box indicates the weight the cohort received (based on inverse variance weighting). The pooled estimate for all cohorts is represented by a black diamond, with the outer edges of the diamond indicating the confidence interval limits.

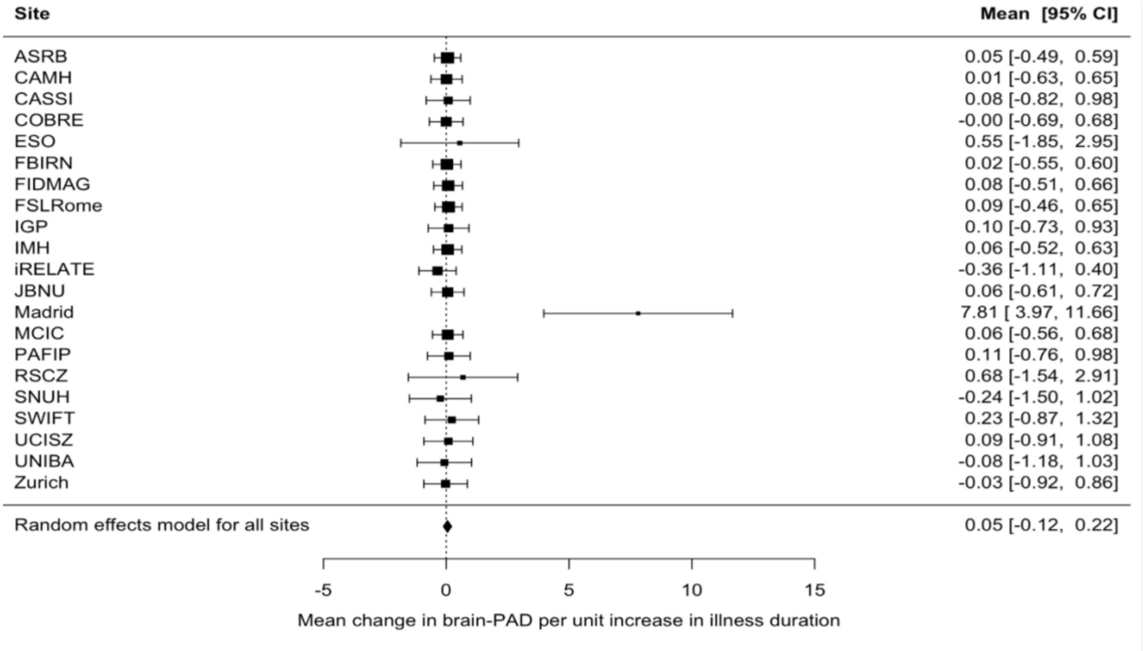

**Supplementary Figure S5b.** Duration of illness and brain-PAD. Forest plot of the association between illness duration and brain-PAD in SZ across 21 cohorts (N=2056). Regression coefficients are denoted by black boxes and indicate a mean change in brain-PAD (years) per unit increase in duration of illness (years), adjusting for age and age$^2$. Black lines indicate 95% confidence intervals. The size of the box indicates the weight the cohort received (based on inverse variance weighting). The pooled estimate for all cohorts is represented by a black diamond, with the outer edges of the diamond indicating the confidence interval limits.

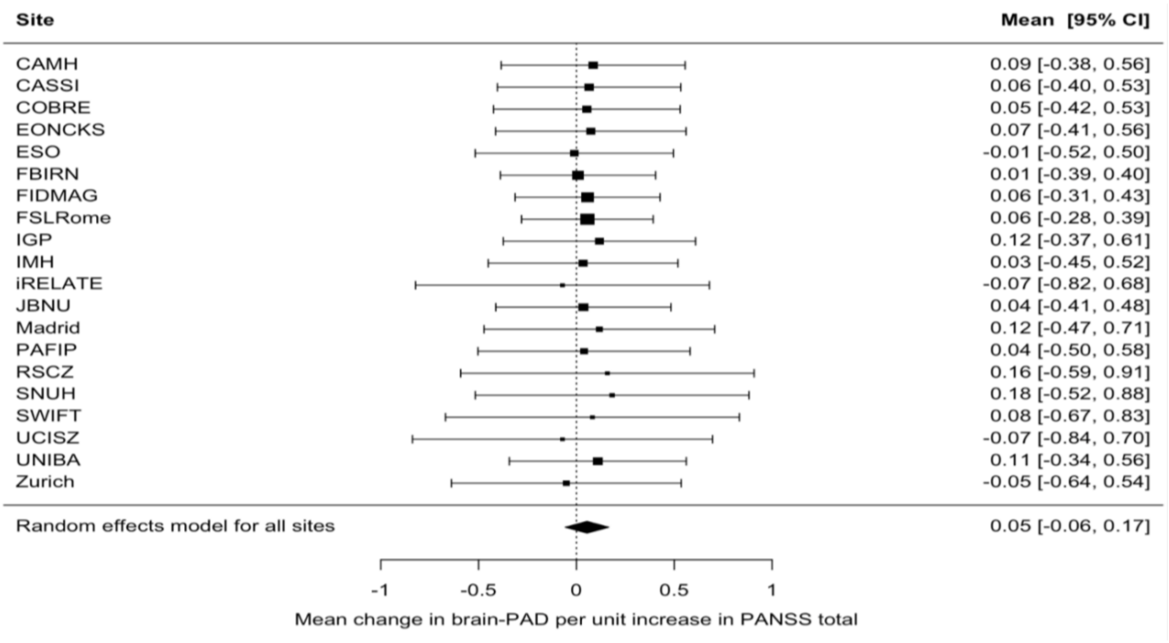

**Supplementary Figure S5c.** Overall illness severity and brain-PAD**.** Forest plot of the association between overall illness severity (PANSS total) and brain-PAD in SZ across 20 cohorts (N=1437). Regression coefficients are denoted by black boxes and indicate a mean change in brain-PAD (years) per unit increase in PANSS total score, adjusting for age and age$^2$. Black lines indicate 95% confidence intervals. The size of the box indicates the weight the cohort received (based on inverse variance weighting). The pooled estimate for all cohorts is represented by a black diamond, with the outer edges of the diamond indicating the confidence interval limits.

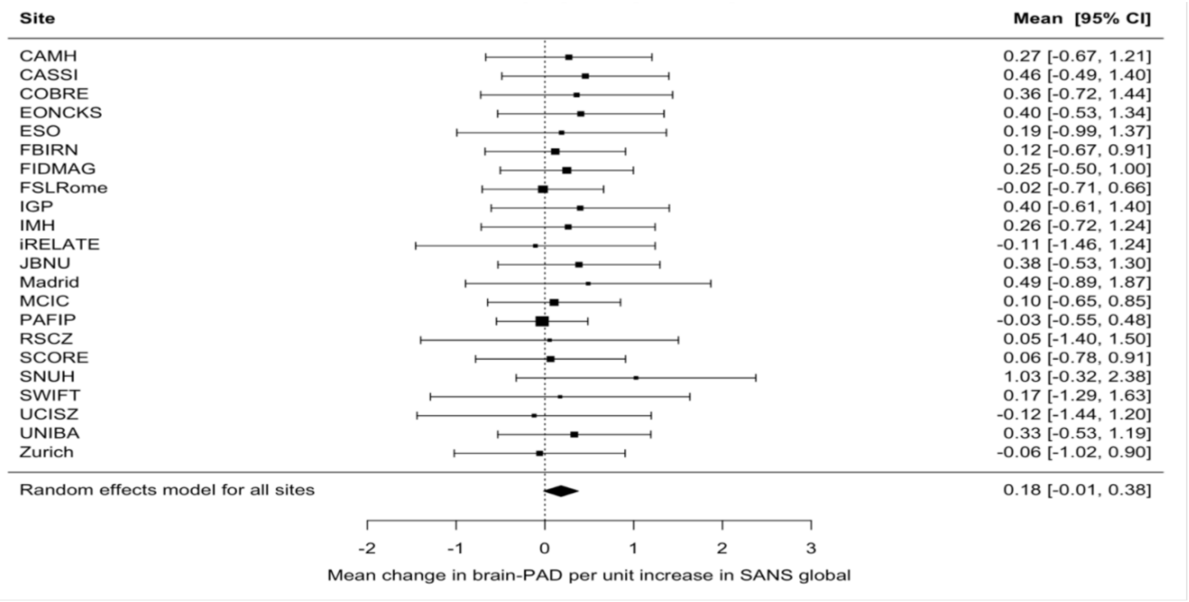

**Supplementary Figure S5d.** Negative symptom severity and brain-PAD. Forest plot of the association between negative symptom severity (SANS global) and brain-PAD in SZ across 22 cohorts (N=1911). Regression coefficients are denoted by black boxes and indicate a mean change in brain-PAD (years) per unit increase in SANS global score, after adjusting for age and age$^2$. Black lines indicate 95% confidence intervals. The size of the box indicates the weight the cohort received (based on inverse variance weighting). The pooled estimate for all cohorts is represented by a black diamond, with the outer edges of the diamond indicating the confidence interval limits.

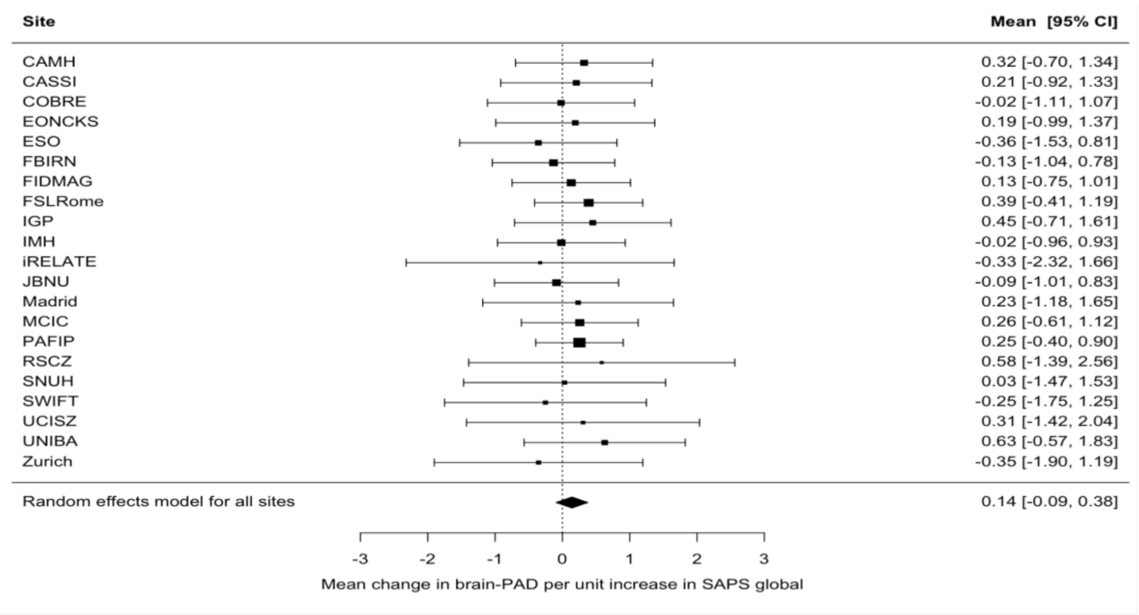

**Supplementary Figure S5e.** Positive symptom severity and brain-PAD. Forest plot of the association between positive symptom severity (SAPS global) and brain-PAD in SZ across 22 cohorts (N=1892). Regression coefficients are denoted by black boxes and indicate a mean change in brain-PAD (years) per unit increase SAPS global score, after adjusting for age and age$^2$. Black lines indicate 95% confidence intervals. The size of the box indicates the weight the cohort received (based on inverse variance weighting). The pooled estimate for all cohorts is represented by a black diamond, with the outer edges of the diamond indicating the confidence interval limits.

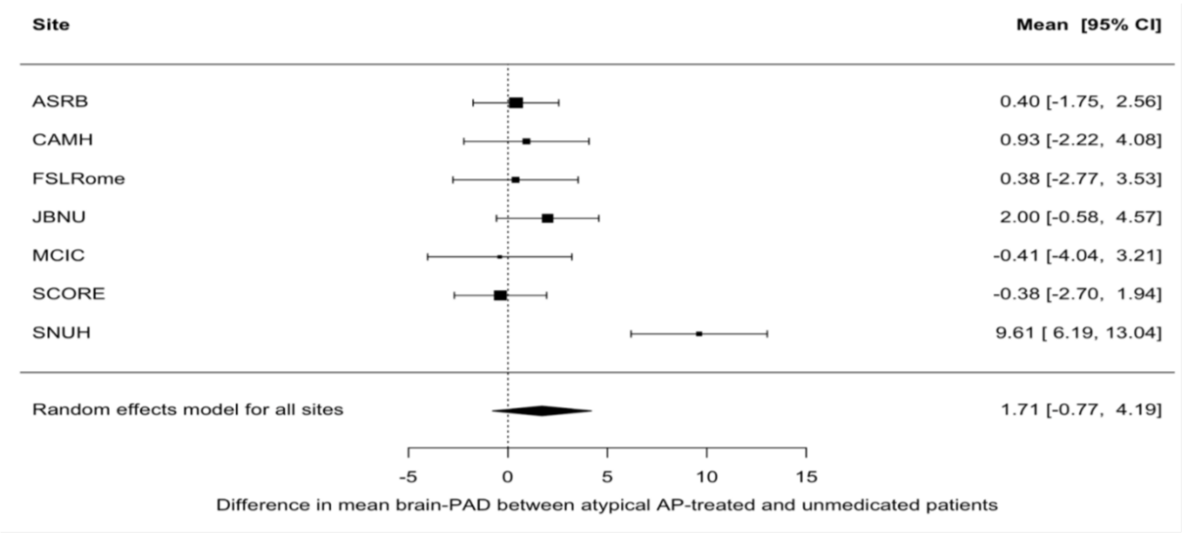

**Supplementary Figure S5f.** Current atypical antipsychotic use and brain-PAD. Forest plot of the association between atypical antipsychotic (AP) use and brain-PAD in SZ across 7 cohorts (N= 642). Regression coefficients are denoted by black boxes and indicate a difference in mean brain-PAD (years) between atypical AP-treated (N=486) and unmedicated patients (N=156), after adjusting for age and age$^2$. Black lines indicate 95% confidence intervals. The size of the box indicates the weight the study received (based on inverse variance weighting). The pooled estimate for all cohorts is represented by a black diamond, with the outer edges of the diamond indicating the confidence interval limits

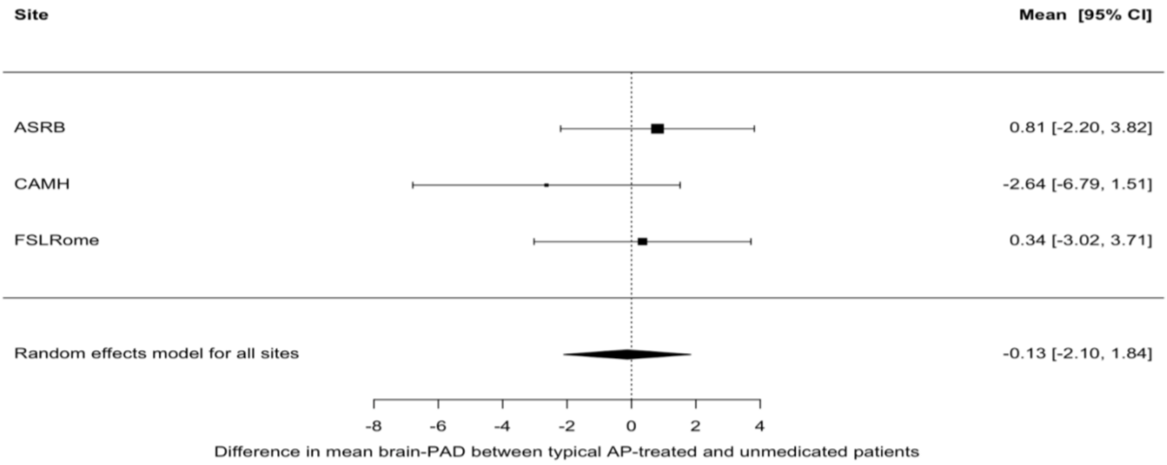

**Supplementary Figure S5g.** Current typical antipsychotic use and brain-PAD. Forest plot of the association between typical antipsychotic (AP) and brain-PAD in SZ across 3 cohorts (N=117). Regression coefficients are denoted by black boxes and indicate a difference in mean brain-PAD (years) between typical AP-treated (N=42) and unmedicated patients (N=72), adjusting for age and age$^2$. Black lines indicate 95% confidence intervals. The size of the box indicates the weight the study received (based on inverse variance weighting). The pooled estimate for all cohorts is represented by a black diamond, with the outer edges of the diamond indicating the confidence interval limits

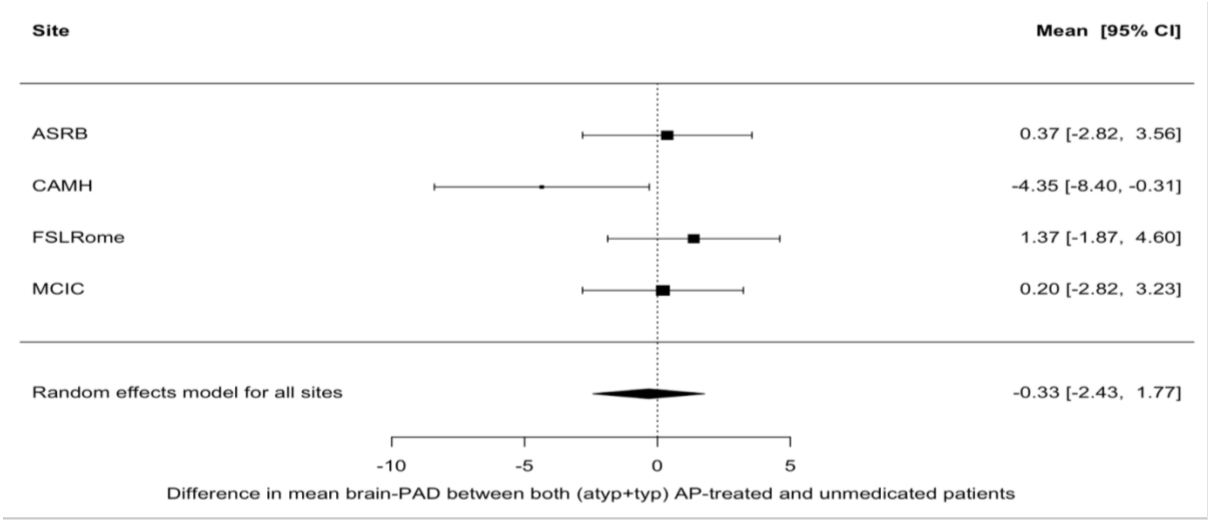

**Supplementary Figure 5h.** Current typical & atypical antipsychotic use and brain-PAD. Forest plot of the association between both typical & atypical antipsychotic (AP) use and brain-PAD in SZ across 4 cohorts (N=266). Regression coefficients are denoted by black boxes and indicate a difference in mean brain-PAD (years) between both (atypical & typical) AP-treated (N=184) and undedicated patients (N=82). Black lines indicate 95% confidence intervals. The size of the box indicates the weight the cohort received (based on inverse variance weighting). The pooled estimate for all cohortsis represented by a black diamond, with the outer edges of the diamond indicating the confidence interval limits

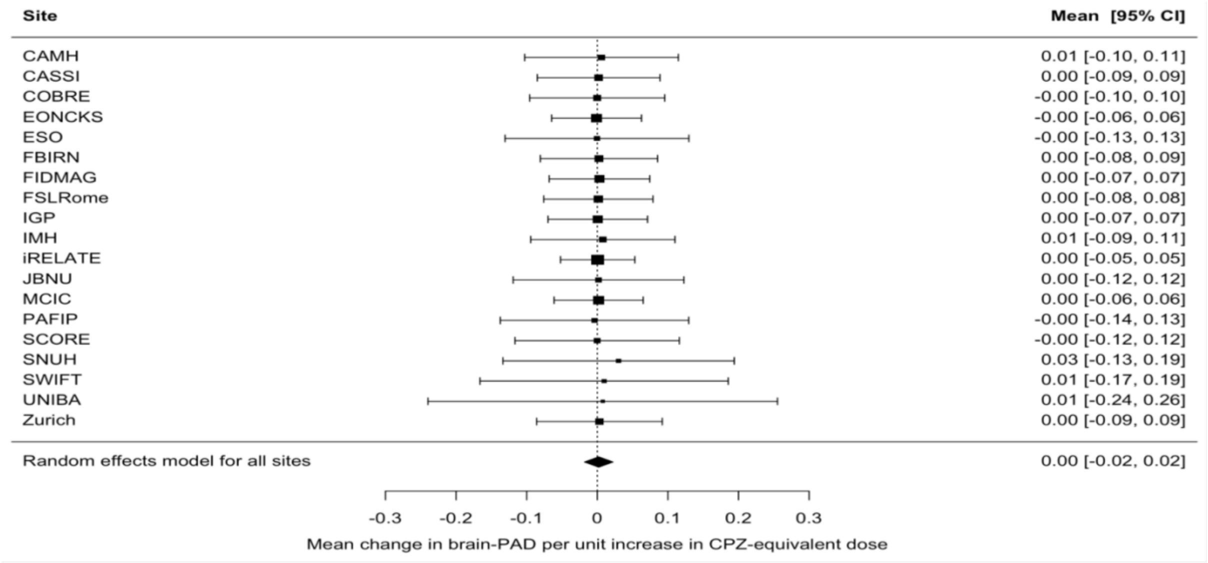

**Supplementary Figure 5i.** Antipsychotic daily dose and brain-PAD. Forest plot of the association between antipsychotic daily (CPZ-equivalent) dose and brain-PAD in SZ across 19 cohorts (N=1698). Regression coefficients are denoted by black boxes and indicate a mean change in brain-PAD (years) per unit increase CPZ-equivalent dose (mg per day), after adjusting for age and age$^2$. Black lines indicate 95% confidence intervals. The size of the box indicates the weight the cohort received (based on inverse variance weighting). The pooled estimate for all cohorts is represented by a black diamond, with the outer edges of the diamond indicating the confidence interval limits
