## Supplementary Material for "Brain ageing in schizophrenia: evidence from 26 international cohorts via the ENIGMA Schizophrenia consortium"

### Image exclusion criteria

As described in Han *et al.* [1], a neuroimaging expert at each scanning site inspected each image segmentation by overlaying the segmentation label of each structure on the T1-weighted brain scan. Additionally, study-wide statistics were collected (means and standard deviations) as well as histogram plots to identify non-normally distributed data and major outliers. Samples were marked if its FreeSurfer feature was >2.698 standard deviations away from the global mean. If a sample was marked as a statistical outlier, the individual site was asked to re-inspect the subject's segmentation to verify that it was properly segmented. If a sample was a statistical outlier, yet properly segmented, it was kept in the dataset. Otherwise, the sample was removed.

### Quality checking and sample exclusion criteria

For each of the participating cohorts, we excluded those participants below 18 years and above 75 years old (if any) to focus on adult patients and controls, and to match the age range of the training dataset previously used to develop the ENIGMA brain age prediction model (see Han *et al*. [1] for further details on the rationale for this age range). We also checked individual FreeSurfer features for missing values and excluded participant samples with >10% missing data, suggestive of poor reliability. In addition, following calculation of brain-PAD for each participant, we intended to winsorize extreme brain-PAD outliers - here defined as data points outside the 5*interquartile range (IQR) within each cohort. However, we identified no outliers outside the 5*IQR at each cohort.

Cohorts with less than 5 participants per group with respect to diagnosis (SZ/HC) or sex (males/females) were excluded from subsequent analyses. As a result of this criterion, one cohort was excluded from the primary case-control analysis due to a HC sample size of less than 5, but SZ data from this site (n=11) contributed to additional analyses (i.e., within the SZ group). Note that for any given clinical characteristic assessed (age of onset, length of illness, symptom severity scores, and antipsychotic use and dose), some of the participating cohorts had no data available (see Supplementary Table S2 for more details) and thus were automatically excluded from additional analyses. In addition, cohorts with available data on less than 5 SZ patients per clinical characteristic (or subgroups for antipsychotic use) were excluded. This resulted in 6 cohorts being excluded from additional analyses on the effects of antipsychotic use.

**Data harmonization for clinical characteristics**

Symptom severity was assessed using the Scale for the Assessment of Negative Symptoms (SANS) [2], the Scale for the Assessment of Positive Symptoms (SAPS) [3], and the Positive and Negative Syndrome Scale (PANSS) [4]. To harmonise symptom severity scores, we decided to convert all positive (i.e., PANSS-Positive and total SAPS composite scores) and negative (i.e., PANSS-Negative and total SANS composite scores) to Global SAPS and Global SANS (summary) scores, respectively. These conversions were made based on recommendations by Andersen [2, 3] and using the algorithms published in van Erp *et al*. [5]. In addition, data on medication dose for typical and atypical antipsychotics were harmonised based on chlorpromazine (CPZ) dose equivalents, as described by Woods (www.scottwilliamwoods.com/files/%0DEquivtext.doc)

**Model validation**

As described in Han *et al*. [1], tenfold cross-validation was performed in the training sample of healthy controls (from the ENIGMA MDD working group) to assess model performance. To quantify model performance, we calculated the (1) mean absolute error (MAE) between predicted brain age and chronological age, (2) Pearson correlation coefficients between predicted brain age and chronological age(r), and (3) the proportion of the variance explained by the model ($R^2$). The accuracy of the model was further validated using a hold-out dataset of healthy controls from the same scanning sites as the training dataset (males, N = 927; females, N = 1199). To evaluate generalizability of the model, independent control test samples (acquired on completely independent scanning sites) from the ENIGMA-BD working group were used.

Training model parameters were applied on these independent subjects (males, N = 610; females, N = 720) from the ENIGMA BD working group.

Within the training set of controls, under cross-validation, the structural brain measures predicted chronological age with a MAE of 6.32 (SD 5.06) years in males and 6.59 (5.14) years in females. Correlation between chronological age and predicted brain age in the cross-validation training sample was r = 0.85, p < 0.001 for both males and females (both $R^2$ = 0.72). Model performance in the hold-out dataset (males MAE = 6.50 [SD 4.91]; r = 0.85, p < 0.001; $R^2$ = 0.72 and females MAE = 6.84 [5.32]; r = 0.72, p < 0.001; $R^2$ = 0.69) was comparable to that of the cross-validation training sample. When applying the model parameters to the independent healthy control samples of the ENIGMA BD working group the generalization worked fairly well (MAE = 7.49 [SD 5.89]; r = 0.71, p < 0.001; $R^2$ = 0.45 for males and MAE = 7.26 [5.63]; r = 0.72, p < 0.001; $R^2$ = 0.48, for females) (for more details see Han *et al*. [1]).

**Correcting for the systemic age bias in brain-age prediction in subsequent analyses**

“Regression dilution” is a well-known phenomenon in any brain age prediction framework, which often leads to young people being systematically predicted to be of older age (than their chronological one) and older people to be systematically predicted younger than their chronological age [6–9]. To account for this potential bias, we have included chronological age as a covariate in subsequent analyses, as proposed elsewhere [6]. This removes all linear age effects on our outcome variable (brain-PAD). However, one cannot assume that the effect of aging on imaging measures is perfectly linear across the lifespan [7, 10, 11]. Therefore, to control for potential non-linear age effects on brain age estimation and brain-PAD, we also included quadratic age ($age^2$) in our subsequent analyses, as suggested elsewhere [7].

**Feature importance: correlations between brain imaging features and brain age**

All imaging features, except the mean lateral ventricle volume, were negatively correlated with predicted brain age (and are visualised in **Figure 2** in the main manuscript), although thickness features correlated more strongly with brain age (mean Pearson r [SD]: −0.46 [0.13]) than subcortical volumes (−0.32 [0.30]) or surface area features (−0.22 [0.06]). We also visualized these associations separately for controls and SZ patients with similar results, suggesting comparable structure coefficients in both groups (**Supplementary Figure S6;** see below). With regards to spatial patterns of brain ageing, our correlation analysis of individual FreeSurfer measures and brain-predicted age does not allow for a straightforward interpretation of the importance of the specific brain regions contributing to the ageing pattern (i.e., we did not include a spatial weight map of our brain age model, as the weights were obtained from a multivariate model). Nevertheless, our results indicate that the ENIGMA brain age model used here relied most heavily on cortical thickness features for making predictions - consistent with the results of its first report by Han *et al*. [1]. The widespread and relatively strong negative correlations between cortical thickness and brain age (as observed here) are in alignment with the general pattern of greater cortical thinning with advancing age reported in the existing literature [12].

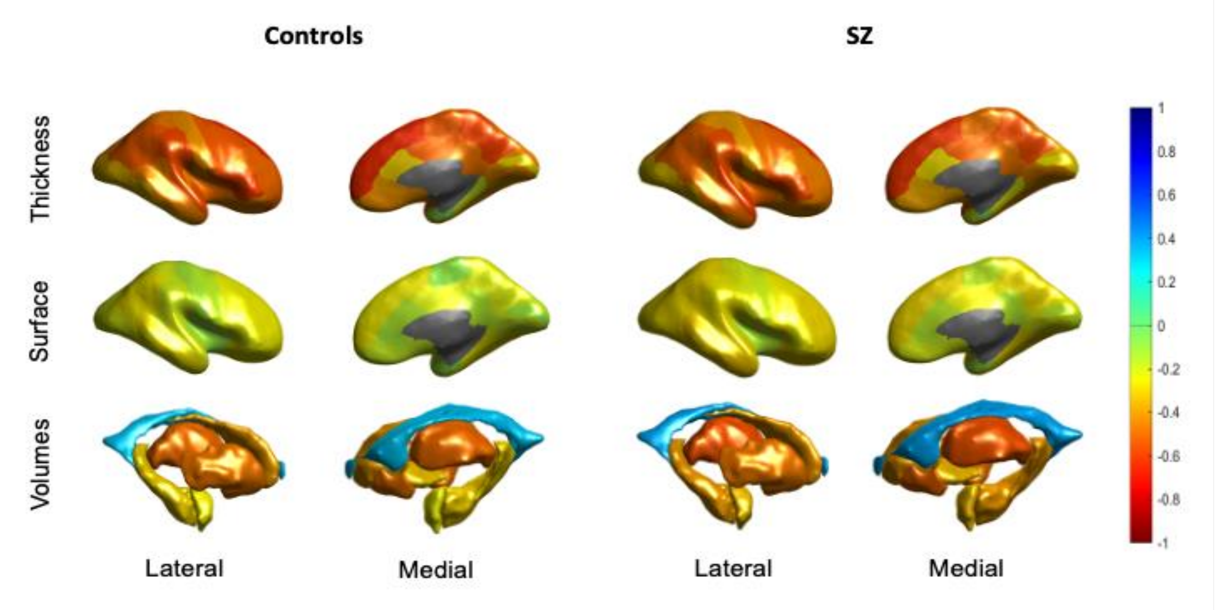


**Supplementary Figure S6.** Correlation coefficients of predicted brain age and FreeSurfer features in healthy control (left) and schizophrenia (right) groups across participating cohorts**.** Bivariate correlations are shown for illustrative purposes to provide a sense of the relative contribution of features in brain age prediction and to show the similarity of patterns between controls and SZ patients. The figure shows Pearson correlations between predicted brain age and cortical thickness features (top row), cortical surface areas (middle row) and subcortical volumes (bottom row), from both the lateral (left) and medial (right) view. The negative correlation with ICV was excluded from this figure for display purposes.
